# PrP concentration in the central nervous system: regional variability, genotypic effects, and pharmacodynamic impact

**DOI:** 10.1101/2021.11.01.21265619

**Authors:** Meredith A Mortberg, Hien T Zhao, Andrew G Reidenbach, Juliana E Gentile, Eric Kuhn, Jill O’Moore, Patrick M Dooley, Theresa R Connors, Curt Mazur, Shona W Allen, Bianca A Trombetta, Alison McManus, Matthew R Moore, Jiewu Liu, Deborah E Cabin, Holly B Kordasiewicz, Joel Mathews, Steven E Arnold, Sonia M Vallabh, Eric Vallabh Minikel

## Abstract

Prion protein (PrP) concentration controls the kinetics of prion replication and is a genetically and pharmacologically validated therapeutic target for prion disease. In order to evaluate PrP concentration as a pharmacodynamic biomarker and assess its contribution to known prion disease risk factors, we developed and validated a plate-based immunoassay reactive for PrP across six species of interest and applicable to brain and cerebrospinal fluid (CSF). PrP concentration varies dramatically between different brain regions in mice, cynomolgus macaques, and humans. PrP expression does not appear to contribute to the known risk factors of age, sex, or common *PRNP* genetic variants. CSF PrP is lowered in the presence of rare pathogenic *PRNP* variants, with heterozygous carriers of P102L displaying 55% and of D178N just 31% the CSF PrP concentration of mutation-negative controls. In rodents, pharmacologic reduction of brain *Prnp* RNA is reflected in brain parenchyma PrP, and in turn in CSF PrP, validating CSF as a sampling compartment for the effect of PrP-lowering therapy. Our findings support the use of CSF PrP as a pharmacodynamic biomarker for PrP-lowering drugs, and suggest that relative reduction from individual baseline CSF PrP concentration may be an appropriate marker for target engagement.

## Introduction

Prion disease is a fatal neurodegenerative disease caused by misfolding of the prion protein (PrP) leading to a gain of toxic function^1^. Lowering PrP expression in the brain is a potential therapeutic approach thoroughly underpinned by genetic proofs of concept^2,3^. Antisense oligonucleotides that lower PrP extend survival by up to three-fold in prion-infected mice^4–6^, supporting the further development of PrP-lowering drugs. This motivates a need to accurately measure the degree to which PrP has been lowered upon drug treatment, across a variety of species and matrices. Such quantification of target engagement — a drug’s impact on its intended molecular target — is critical throughout the life cycle of any drug development program, from therapeutic candidate screening and lead optimization, to in vitro and in vivo pharmacology studies in animals, to dose selection and confirmation of drug activity in human clinical trials. In prion disease, quantification of PrP may play an even larger role: lowering of cerebrospinal fluid (CSF) PrP in presymptomatic individuals at high risk for genetic prion disease could be employed as a surrogate biomarker endpoint in support of provisional drug approval^3^.

In previous studies, PrP in human CSF has been quantified using a commercially available ELISA assay specific to human PrP^7–11^, as well as a multiple reaction monitoring (MRM) targeted mass spectrometry assay^12^. PrP is highly abundant in human CSF^12^, on the order of tens or hundreds of nanograms per milliliter. CSF PrP concentration paradoxically decreases in symptomatic prion disease^7–10,12,13^ amidst a toxic buildup of PrP in the brain, but no decline in CSF PrP was observed in presymptomatic mutation carriers^11^. PrP sticks to plastic, and is thus exquisitely sensitive to preanalytical variables, but with uniform sample handling and addition of detergent, CSF PrP can be reliably quantified^10^, with a test-retest mean coefficient of variation (CV) of only 7% in serial samples collected from the same individuals over more than a year^11^. These findings support the use of CSF PrP as a pharmacodynamic biomarker to measure target engagement in presymptomatic individuals.

Despite this strong foundation, the development path for PrP-lowering therapeutics faces several outstanding practical needs, including improved measurement tools both to track treatment response and to better address unresolved biological questions about disease pathophysiology. Drug development activities will be facilitated by establishment of an inexpensive, easy-to-implement assay capable of measuring PrP both in humans and across relevant animal species, in both brain and CSF. Advanced age, male sex, and both rare and common *PRNP* genetic variants are risk factors for prion disease^14,15^, and it is unknown whether differences in PrP expression contribute to any of these factors. Regional differences in brain PrP expression^16–18^ might interact with drug distribution patterns in the brain^19^ to influence biomarker and clinical outcomes in future trials. Expectations that pharmacologic lowering of PrP RNA in the brain should be reflected in brain PrP and consequently in CSF PrP should be experimentally demonstrated in animals to validate the use of CSF as a sampling compartment. Here, we develop a new cross-species PrP ELISA assay, assess its performance characteristics, and deploy it across a range of animal and human samples to address the above questions.

## Results

### Cross-species ELISA assay

After screening four commercially available anti-PrP monoclonal antibodies in pairs for sensitivity and cross-reactivity (Figure S1), we developed a final assay protocol (Appendices 1-2) using monoclonals EP1802Y for capture and 8H4 for detection, with C-terminal epitopes respectively mapped to approximately residues 218-227 and 182-196 (human codon numbering)^20–22^. The assay possesses dynamic range from 0.05 to 5.0 ng/mL, exhibits linearity for endogenous PrP in mouse brain homogenate, and meets FDA criteria for bioanalytical method validation^23^ (Table S1 and S2), except for elevated inter-plate variability near the lower limit of quantification (Table S1). Quantification of PrP in brain homogenate required 0.2% CHAPS to fully solubilize PrP (Figure S1), minimization of time spent above freezing (Table S2), and plating at uniform dilution (Figure S2). The assay is applicable to both brain and CSF and is equally reactive with human, cynomolgus, mouse, rat, and bank vole PrP, with slightly reduced reactivity for Syrian hamster PrP (Figure S2, S3, S4, Table S3).

### Regional distribution of brain PrP

We previously observed an ∼8-fold difference in PrP concentrations among N=28 human brain samples^10^, which could have reflected one or more of the following: brain region differences, inter-individual differences, effects of agonal state or post-mortem interval, and/or, preanalytical variation due to incomplete solubilization of PrP in the 0.03% CHAPS buffer used at that time. We therefore obtained a new set of 6-7 matched brain regions from each of 5 control individuals, and homogenized them in 0.2% CHAPS for analysis by cross-species PrP ELISA. We identified considerable regional differences (*P* = 0.0007, Type I ANOVA), with PrP almost ten times higher in parietal cortex (BA7) than in olivary nuclei (Figure 1A-B). Analogous regional disparities were observed in cynomolgus macaques (*P* < 1e-10, Type I ANOVA, Figure 1C-D) and mice (*P* < 1e-10, Type I ANOVA, Figure 1E-F). Across all three species, PrP concentrations were higher in cortex than in subcortex and cerebellum, and were lowest in brainstem, although humans displayed the highest inter-individual variability and a low PrP concentration in hippocampus in contrast to mouse and macaque (see Discussion).

**Figure 1.**
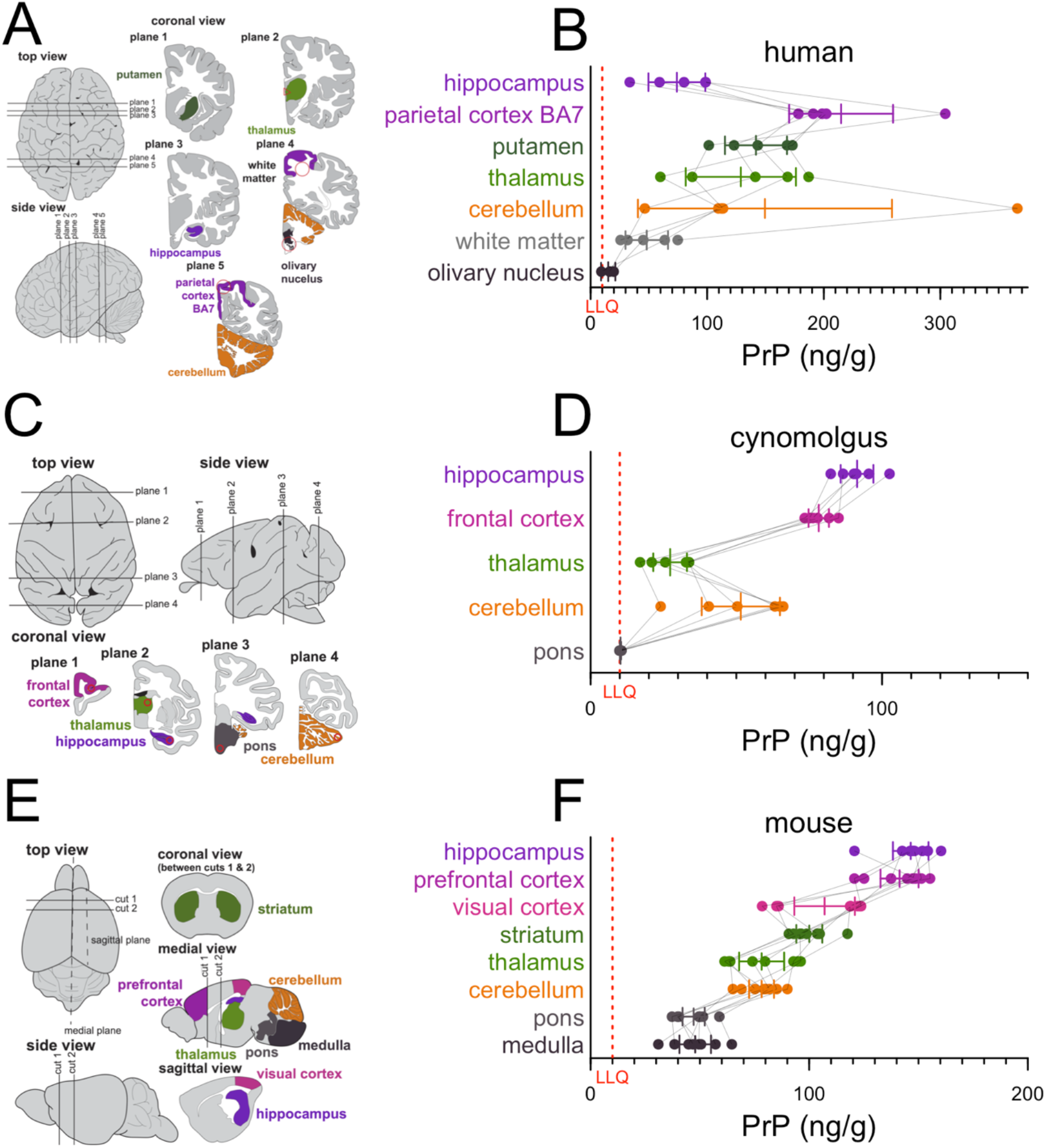
Regional distribution of brain PrP. **A, C, E)** Diagrams of brain regions examined in humans, cynomolgus macaques, and mice, and. **B, D, F)** PrP concentrations in N=5 human, N=6 macaque, and N=8 mouse brains. Thin lines connect regions from the same individual. Bars indicate mean and 95% confidence interval of the mean. Red dashed lines indicate lower limit of quantification (LLQ). Brain diagrams were traced from Allen Brain Atlas images^24,25^.

### Assessment of sex and age effects on PrP expression

We first analyzed *PRNP* RNA levels in the GTEx v8 dataset^26^. After controlling for cause of death (4-point Hardy scale), which is confounded with sex and with decade of life (*P* < 1e-10 for both, Chi-squared test), and correcting for multiple testing, only minor salivary gland and skeletal muscle showed any evidence of age-dependent expression (higher with age, Figure 2A), and only mammary tissue and cultured fibroblasts showed evidence of sex-biased expression (higher in females, Figure 2B). We found no evidence that *PRNP* RNA expression in any brain region (yellow, Figure 2A-B) correlated with age or sex. PrP protein expression might nevertheless change in brain parenchyma due to changes in translation or degradation rates, however, considering differences in PrP concentration across brain regions found here (Figure 1) and by others^16–18^, and the potential impact of preanalytical variables (Table S2), we were unable to identify a sample set of human brains suitable for querying age differences. We therefore confined our subsequent analyses to human CSF and to rat brain and CSF.

**Figure 2.**
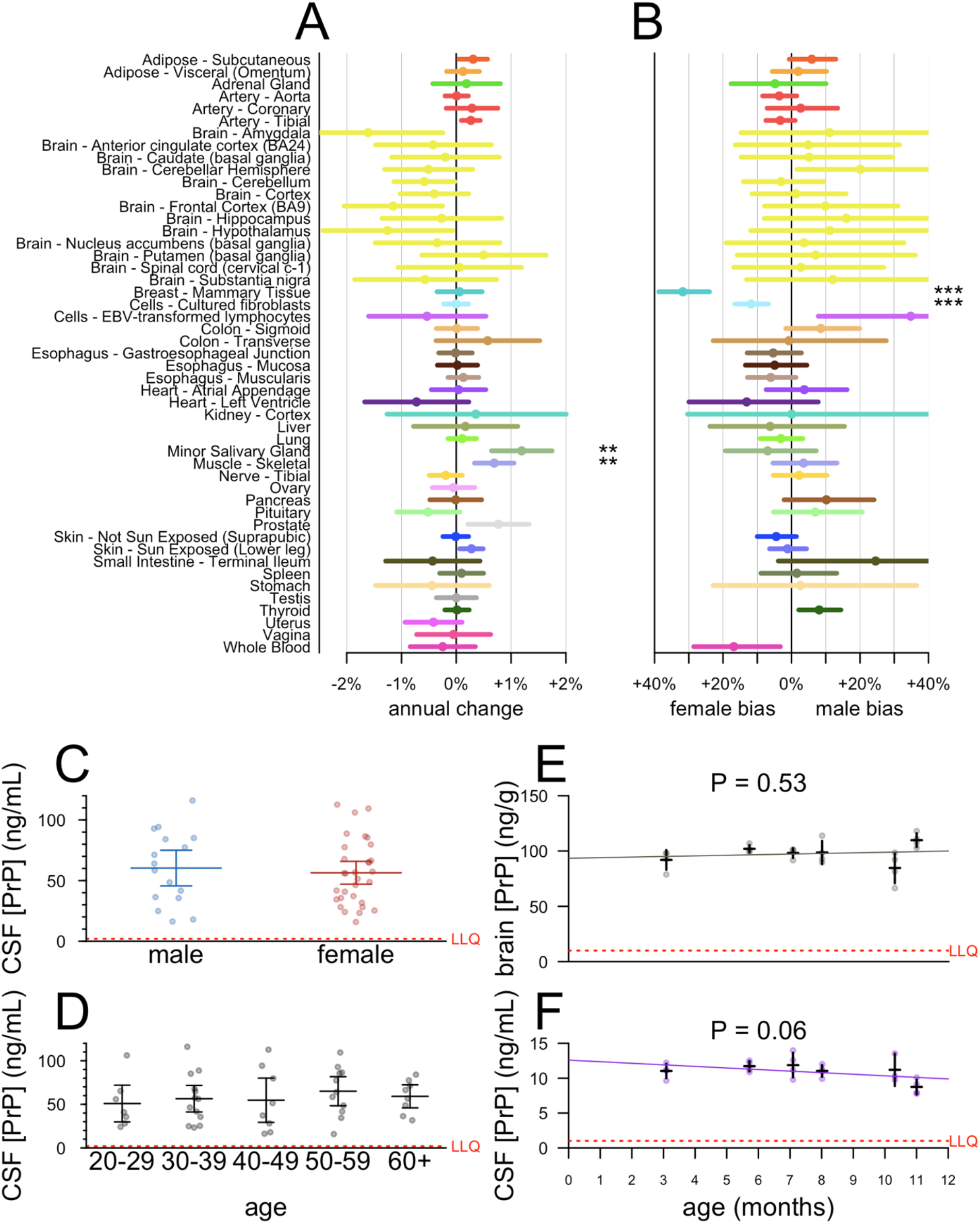
Lack of evidence for sex or age effects in PrP expression. **A-B)** Analysis of publicly available GTEx v8 data. Log-linear models log(tpm) ∼ age + hardy + sex (see Results text) were fit for each tissue, and the mean annual change (dots) was calculated as exp(beta_age_)-1 and exp(beta_sex_) respectively, with 95% confidence intervals (line segments) given by 1.96 standard errors of the mean. After Bonferroni correction for N=49 tests (A) or N=44 tests (B), symbols indicate * P < 0.05, ** P < 0.01, *** P < 0.001. **C-D)** CSF PrP concentrations averaged across all available CSF samples for N=47 MGH study participants stratified by sex (C) or age (D). Bars indicate mean and 95% confidence interval of the mean. **E-F)** brain (E) and CSF (F) concentrations of PrP for cohorts of N=4 male Sprague-Dawley rats age 3-11 months. Red dashed lines indicate lower limit of quantification (LLQ).

We measured PrP in CSF from N=47 individuals (healthy asymptomatic *PRNP* mutation carriers and non-carrier controls) from our cohort study at Massachusetts General Hospital (MGH)^11^. Exquisite uniformity of CSF handling plus early addition of 0.03% CHAPS minimize preanalytical confounders in this cohort. Among the N=36 of these individuals who had >1 serial sample (range: 2-5 lumbar punctures performed over a period up to 3.5 years), CSF PrP measured in cross-species ELISA exhibited tight test-retest reliability (mean CV=11.1%). We therefore focused on each individual’s mean CSF PrP value observed across all visits. We found no evidence for CSF PrP association with sex (*P* = 0.81, Kolmogorov-Smirnov test, Figure 2C), nor age (P = 0.28, Spearman correlation, Figure 2D), nor. In male rats aged 3-11 months, PrP concentrations in neither brain (Figure 2E) nor CSF (Figure 2F) changed with age.

### Genotypic effects on human CSF PrP concentration

In N=47 cohort study participants with at least one CSF sample available, we examined genotypic differences in CSF PrP by comparing mean CSF PrP levels averaged, for each individual, across all study visits (Figure 3A). Compared to mutation-negative controls (N=21), CSF PrP was lower for carriers of P102L (55%, P = 0.0055, Kolmogorov-Smirnov test, N=4) and D178N (31%, P=6.7e-6, Kolmogorov-Smirnov test, N=6); the trend was preserved but non-significant for E200K (78%, P = 0.23, Kolmogorov-Smirnov test, N=12).

**Figure 3.**
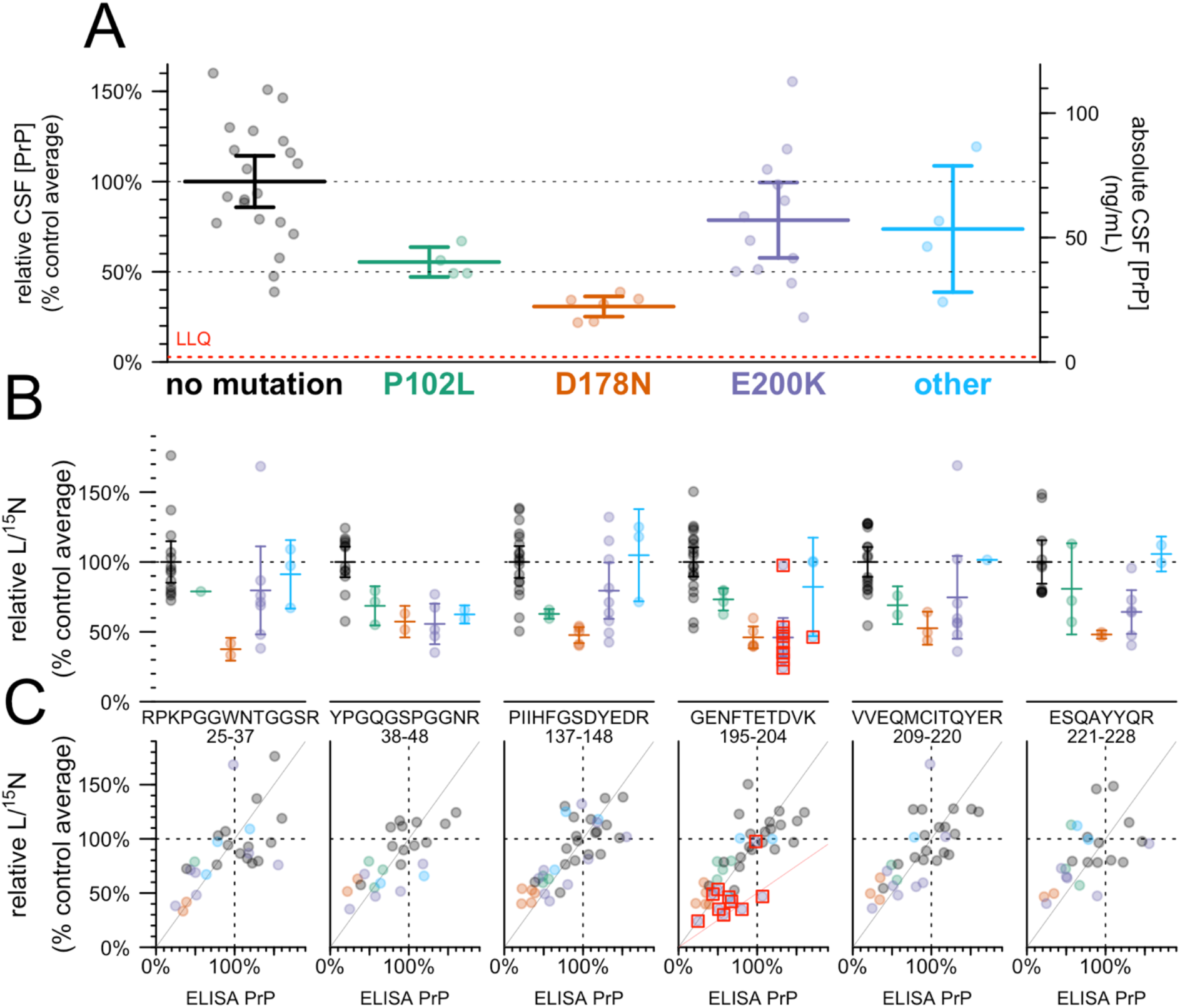
Effect of PRNP mutation on CSF PrP concentration. **A)** CSF PrP concentrations measured by cross-species ELISA, averaged across all available CSF samples for each of N=47 MGH study participants, normalized to the mean of non-mutation carrier controls. Bars indicate mean and 95% confidence interval of the mean. Red dashed line indicates lower limit of quantification (LLQ). This sample set includes N=29 individuals for which CSF PrP concentrations determined by BetaPrion ELISA were previously reported^11^. **B)** The same samples analyzed by the PrP MRM assay, peptides arranged from N-terminal (left) to C-terminal (right). Peptide sequences and residue numbers are noted beneath each plot. Note that because samples where technical replicates had CV > 15% are removed, the number of samples differs for each panel. Bars indicate mean and 95% confidence interval of the mean. **C)** Correlation between ELISA results from A (x axis) and MRM results from B (y axis), with lines indicating a diagonal with slope = 1 (gray) and 0.5 (pink, GENFTETDVK only). In B and C, red boxes indicate individuals whose mutation abolishes the tryptic peptide being monitored in that plot.

ELISA relies on the presence of two intact epitopes on the same protein, so non-reactivity of one of our antibodies for one of these mutations could give rise to artifactual genotypic differences. The 8H4 epitope has been mapped to a region adjacent to D178N and E200K^20,21^, and some PrP mutations are reported to affect interdomain interactions^27^ and could therefore alter the accessibility even of distal epitopes^21^. We therefore employed a targeted mass spectrometry method^12^ using stable isotope labeled amino acids and MRM, on CSF, to measure six tryptic peptides spanning the N to C terminus of PrP. Individuals with the E200K, P102L, and particularly D178N mutations, had lower mean levels of all six peptides, compared to mutation-negative controls (Figure 3B). Indeed, across individual samples, those that were low in ELISA were low in MRM and those that were high in ELISA were high in MRM, with samples clustering along the diagonal with a slope equal to one (gray line, Figure 3C). For peptide GENFTETDVK, those individuals whose mutations disrupt this peptide (mostly E200K individuals; red boxes, Figure 3C) clustered closer to a line with slope equal to 0.5 (pink line, Figure 3C), consistent with non-detection of this peptide from the mutant allele. Overall, the fact that each peptide observed in MRM replicates the ELISA result confirms that CSF PrP is genuinely reduced in a genotype-dependent manner in individuals with certain pathogenic *PRNP* mutations.

We also examined two common variants in *PRNP*: M129V (rs1799990), and a non-coding variant 72 kb upstream of *PRNP* (rs17327121) implicated as the lead variant for an expression quantitative trait locus (eQTL) in cerebellum^26^. Neither was significantly correlated with CSF PrP in our samples (Figure S5).

### Pharmacodynamic effect of PrP RNA-targeting therapy in rodents

*Prnp*-targeting ASOs that extend survival in vivo do so by lowering *Prnp* RNA^5,6,28^. This reduction in *Prnp* RNA is expected to lead to lowering of brain parenchyma PrP, and in turn to reduction of PrP released into CSF, but the relationship between these variables has not yet been quantitatively investigated.

We first sought to understand the relationship between whole-brain *Prnp* RNA and protein levels in mice using ASO 6, a tool compound previously shown to extend survival of prion-infected mice^6^. At two and four weeks post-dose in naïve animals, active ASO 6 dose-dependently suppressed whole hemisphere PrP (Figure 4A-B). Protein suppression was weaker than RNA suppression at two weeks, with each 1% reduction in *Prnp* RNA corresponding to just a 0.62% reduction in PrP (Figure 4A). The two measures were in closer agreement by four weeks, with each 1% RNA reduction corresponding to 0.83% PrP reduction (Figure 4B). We observed comparable target engagement and close correspondence between RNA and protein levels in RML prion-infected animals treated at 60 dpi and harvested four weeks post-dose at 88 dpi (Figure 4C).

**Figure 4.**
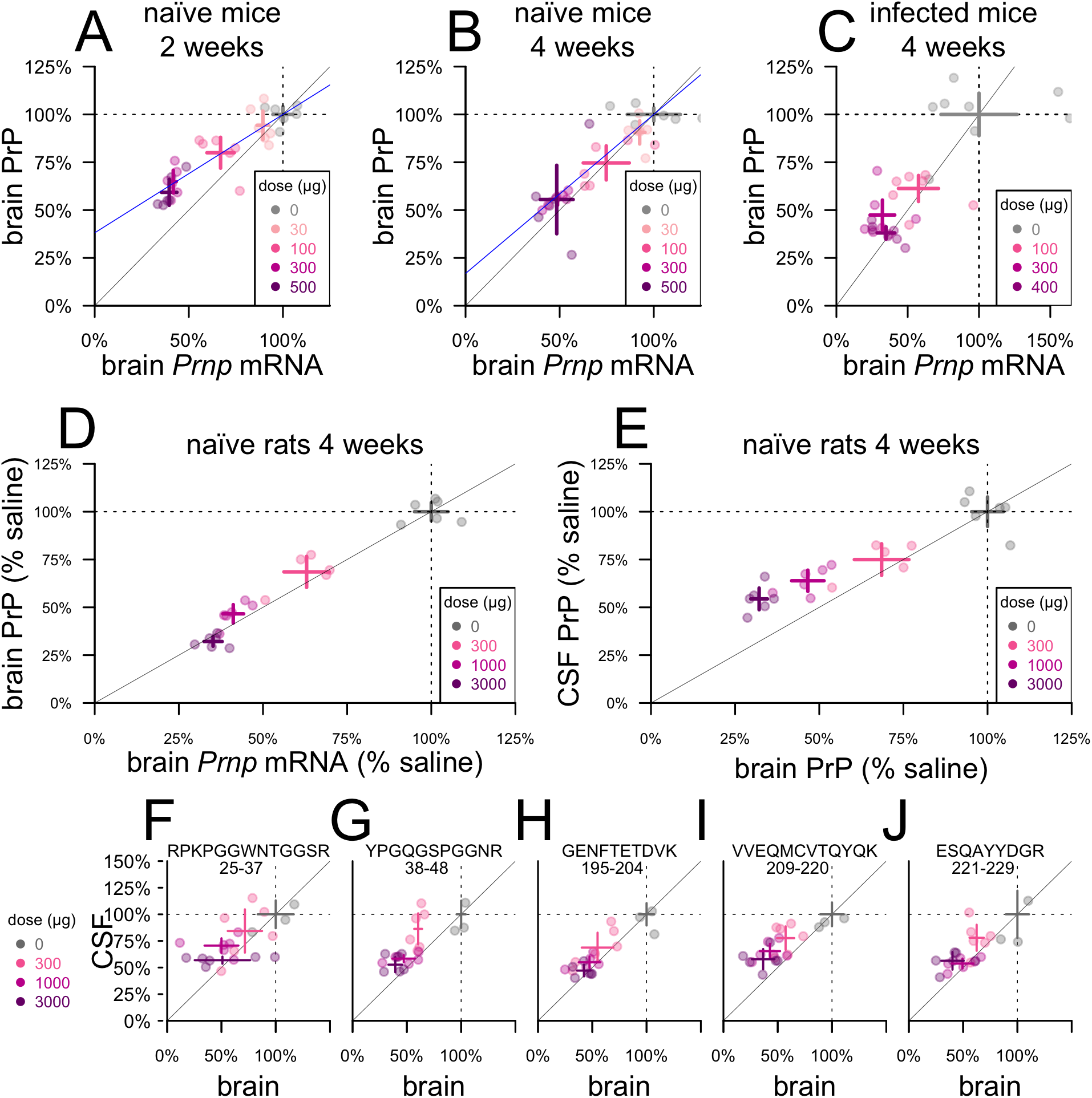
Pharmacodynamic effect of PrP RNA-targeting therapy. **A-B)** Whole-hemisphere RNA (x axis) vs. PrP (y axis) reduction measured by ELISA in groups of N=6 naïve mice at 2 weeks (A) and 4 weeks (B) post-dose. Blue lines represent linear regression best fits with the (1,1) coordinate fixed. C) RNA from the lateral half of one hemisphere (x axis) vs. PrP from the medial half of the same hemisphere (y axis) reduction measured by ELISA in groups of N=6 RML prion-infected mice dosed at 60 dpi and harvested at 4 weeks post-dose. D) Whole-hemisphere RNA (x axis) vs. PrP (y axis) reduction measured by ELISA in groups of N=6 naïve rats harvested at 4 weeks post-dose. E) Whole hemisphere PrP (x axis) reduction vs. CSF PrP (y axis) in the same rats. F-J) CSF and brain samples from panel E analyzed by MRM, with the five rat PrP peptides arranged from N-terminal (F) to C-terminal (J). Peptide sequences and residue numbers are noted above each plot. In every panel, crosshairs represent the mean and 95% confidence interval of the mean on both dimensions.

Because CSF PrP is more sensitive to plastic adsorption when handled in very small volumes^10^, it would be challenging to measure CSF PrP reduction upon drug treatment in mice. We therefore examined the relationship between *Prnp* mRNA, brain PrP, and CSF PrP in rats (Figure 4D-E). At four weeks post-dose, whole hemisphere PrP was dose-dependently suppressed in proportion to whole hemisphere *Prnp* mRNA (Figure 4D). The reduction in brain PrP was in turn reflected in CSF PrP, although CSF PrP reduction slightly underestimated the depth of target engagement in brain parenchyma, with each 1% reduction in CSF PrP knockdown corresponding to a 1.4% reduction in brain PrP (Figure 4E). The relationship between PrP knockdown in brain and in CSF was reproduced by MRM, and did not differ significantly among the five peptides examined (*P* = 0.14, ANCOVA; Figure 4F-J).

## Discussion

Given the pivotal role of PrP in prion biology, it is reasonable to ask whether any known risk factors for prion disease, including age, sex, and genotype, are mediated by differences in PrP expression. Two previous studies observed suggestive associations between CSF PrP concentration and age^10,13^, but only in historical cohorts where preanalytical variables were not well-controlled and/or samples were not well-matched on other variables. One previous study indicated that PrP expression on peripheral leukocytes rose with age^29^, but no such change in brain has been reported beyond the first few weeks of life^17,30^. If PrP expression in brain rose with age, this could potentially explain the mid- to late-life onset of most prion disease, even in the lifelong presence of a pathogenic mutation^31,32^. We found no evidence, however, that human brain *PRNP* RNA expression, PrP concentration in human CSF, or PrP in rat brain and CSF, change with age. If PrP expression were indeed sex-biased, this could potentially explain the reportedly higher incidence of prion disease in men^14^ (risk ratio = 1.2). We found no evidence, however, from publicly available RNA data nor from our own analyses of human CSF, to support a sex difference in PrP expression. Common variants in *PRNP* are associated with prion disease risk, but this risk exhibits no obvious connection to *PRNP* expression^33^. The common variant M129V affects the risk and histopathological subtype of sporadic and acquired prion disease as well as disease duration in genetic prion disease^15,32^, but while it is the lead SNP for a peripheral tissue eQTL, it is not an eQTL in human brain^26^. We found no evidence that M129V affects CSF PrP. The lead variant for a reported cerebellar eQTL 72 kb upstream of *PRNP*^26^, which is not known to be associated to prion disease risk^33^, likewise showed no evidence of influencing CSF PrP. All of the above analyses are underpowered for small effect sizes, but use of larger historical CSF cohorts to interrogate these questions would be complicated by the uncontrolled preanalytical variability present in such samples^10^. While our findings do not rule out sex, age, or common variant effects on PrP expression, they may suggest that any such effects are too small to be major confounders in a clinical trial enrolling tens of individuals.

CSF PrP concentrations are dramatically lower, however, in individuals with some pathogenic *PRNP* mutations. The number of individuals examined here remains small, and includes samples reported previously^11^. Nevertheless, the difference is large and unambiguously statistically significant, and this finding replicates across two ELISA assays^11^ and six peptides monitored by MRM, ruling out an immunoreactivity artifact. This genotypic difference has been maintained over years of follow-up and in the absence of detectable prodromal pathological changes^11^, which appear to occur only very shortly before onset in prion disease^34^. We therefore conclude that these mutations lead to constitutively lower concentrations of PrP in CSF. In principle, this could arise from any combination of the following: reduced translation, faster catabolism, or reduced shedding of PrP into CSF. Studies of D178N in animal and cell culture models favor faster catabolism, perhaps secondary to impaired trafficking, and thus lower steady state levels in parenchyma for this mutation^35–38^. CSF PrP in D178N carriers averaged just 31% that of non-carrier controls. That this number is less than 50%, despite all of our carriers being heterozygotes, raises the possibility that the presence of the mutation suppresses the expression or shedding of the wild-type protein, or shortens its half-life, *in trans*. This possibility, and the mechanism that might govern it, warrant further study.

That CSF PrP differs by genotype prompts consideration of the basis — relative or absolute — on which target engagement should be assessed in clinical trials of PrP-lowering drugs. Clinical trials of ASOs for other targets have generally reported relative reductions in target biomarkers — percentage declines from individual baselines^39,40^. There also exists, however, precedent for therapies being dosed to target an absolute level of a response biomarker. The best predictor of efficacy for the antibody omalizumab in severe asthma is the patient’s free IgE after treatment, with the goal of reducing levels to below 25 ng/mL^41^. Thus, some may ask whether PrP-lowering therapies should be dosed to keep CSF PrP below some absolute ng/mL level. D178N is highly penetrant^42^ and exhibits earlier average onset than E200K^32^ despite reduced basal levels of CSF PrP. This argues that, while CSF PrP appears usable as a therapeutic response marker in prion disease, absolute levels of this analyte may not hold significance that generalizes across individuals. Thus, a single absolute threshold would likely not serve as an appropriate treatment goal.

A proposed pathway^3^ whereby a PrP-lowering drug could receive provisional approval based on lowering of CSF PrP relies crucially on lowering of PrP in brain being reflected in CSF. Here we empirically validate this link in rats, and show that response is uniform across tryptic peptides spanning the length of PrP. Thus, CSF PrP appears to be one analyte, with multiple different measurement methods all reflecting the concentration of the disease-relevant protein.

We found that PrP concentration varies dramatically between different brain regions in humans, in monkeys, and in mice. The pattern was generally consistent across species and agrees with previous reports in rodents^17,18^. Human brain samples exhibited much higher inter-individual variability than monkeys or mice, however, and only modest PrP concentrations were detected in human hippocampal samples, whereas this region had the single highest PrP concentration in both mice and monkeys. These observations might reflect the size of the human brain. For example, whereas we analyzed whole mouse hippocampi, in humans we examined a ∼160 mg medial slice of CA1-4, posterior to the globus pallidus. If PrP expression is highly enriched in certain subregional structures^17,18^, then slight dissection differences could yield varying results. Nonetheless, our findings across species argue that PrP expression exhibits genuinely large variability between regions, and this should be accounted for when modeling which regions contribute to pharmacodynamic signal in CSF.

Our findings also bear on the timescale on which the pharmacodynamic effect of PrP-lowering therapies can be observed. PrP’s half-life in the mouse brain was estimated at just 18 hours in a conditional mouse model^43^, but one PrP peptide detected in the brains of isotopically labeled mice showed a half-life of 5 days^44^. The ASO used here reaches maximal activity at the RNA level within ∼7 days^6^, yet appeared to achieve deeper protein suppression in brain parenchyma at 4 weeks than at 2 weeks post-dose, which would be more consistent with the higher estimate for PrP half-life. We previously observed that following a single ASO treatment in mice, it takes three weeks for neuropathological markers to diverge between treated and untreated cohorts. Levels of plasma neurofilament light, a marker of neuronal damage, continue to decline for six weeks post-dose^6^. These kinetics are consistent with a lag between engagement of the RNA target, reduction of protein levels, and amelioration of the downstream disease process. Together, these findings may inform timing considerations for dosing of PrP-lowering therapies.

Finally, we observed that CSF PrP slightly underestimated the depth of parenchymal PrP knockdown at 4 weeks, which could reflect either a different half-life or different dose-response relationship for PrP released into CSF. More detailed pharmacodynamic modeling in multiple species will be required to link CSF PrP readouts in humans to estimates of brain parenchyma PrP reduction.

Our assay should serve as a tool for further development of PrP-lowering therapies, and our findings support the utility of PrP quantification as a tool in the development paths of such therapies.

## Methods

### Assay development

Initial assay development was undertaken by Bioagilytix Boston (then known as Cambridge Biomedical). Antibody pair and other key assay configuration parameters were established, and the assay was subjected to a full validation study for rat CSF compliant with World Health Organization Good Clinical Laboratory Practice Regulations (GCLP)^45^. The assay was then transferred to the Broad Institute where the standard curve points and reagent concentrations were modified to yield the final assay conditions described below. Validation for mouse brain homogenate and all subsequent studies were performed at the Broad Institute.

### Cross-species PrP ELISA

The exact assay protocol and checklist referred to at the bench while running the assay are provided as Appendices 1 and 2. The method is briefly summarized as follows.

To prepare biotinylated detection antibody, 1 mg of EZ-Link Sulfo-NHS-SS-Biotin (Thermo A39258) was combined with 0.09 mg of 8H4 antibody (Abcam ab61409). Conjugated antibody was purified using Zeba spin desalting columns (Thermo 89889) and quantified by NanoDrop. Assay buffer of 0.05% wt/vol Tween (Teknova T0710), 5% BSA in 1X CSHL PBS was 0.22µm vacuum-filtered and stored at 4°C. Wash buffer was 0.1% Tween in 1X CSHL PBS, stored at room temperature.

Clear flat-bottom MaxiSorp plates (ThermoFisher 439454) were coated overnight at 4°C with 2.0 µg/mL EP1802Y capture antibody (Abcam ab52604) in PBS, sealed with clear adhesive MicroAmp Film (Life 4306311) and then washed 3 times with 300 µL wash buffer and tapped dry (subsequent washes followed this same procedure). Plates were blocked with 250 µL assay buffer (0.05% Tween20, 5% BSA, 1X PBS), sealed at RT for 1-3h and then washed. A fresh aliquot of recombinant PrP was thawed to make a new standard curve for every plate (5, 2, 0.8, 0.32, 0.13, 0.05, and 0 ng/mL). Standards, QCs, and samples were diluted into assay buffer in microcentrifuge tubes and 100 µL was added per well. Plates were sealed and incubated with sample for 60-75 minutes and then washed. Biotinylated 8H4 detection antibody was added at 0.25 µg/mL in 100 µL assay buffer, plates were sealed, incubated 60-75 minutes, and then washed. Pierce High Sensitivity Streptavidin-HRP (Thermo 21130) was added at 24.69 ng/mL in 100 µL assay buffer, plates were sealed, incubated for 30 minutes, and then washed. 100 µL TMB (Cell Signal 7004P4) was added, plates were incubated in darkness but monitored periodically for absorbance at 605 nm. After 30 minutes or when absorbance for the 5 ng/mL standard reached 0.8, whichever came sooner, 100 µL of stop solution (Cell Signal 7002L) was added, plates were shaken briefly and then read at 450 nm with 630 nm background subtraction on a Spectramax M5 platereader (Molecular Devices). Standard curves were fit with a 4-point hill slope curve using the minpack.lm package^46^ in R. FDA guidance^23^ was followed for non-GLP validation of the assay in mouse brain homogenate (Table S1, S2; Figure S1, S2; Appendix 3). To obtain diluted concentrations within the dynamic range of the assay, brains were run at a final dilution of 1:200 (for instance, 1:20 dilution of 10% wt/vol homogenate), while CSF samples were run at dilutions of either 1:20 (rat), or ≥1:40 (human; five samples at or near upper limit of quantification at 1:40 were re-run at 1:80).

For plates prepared in the prion laboratory, the protocol was modified as follows. All reagents, standard curves, and QCs were diluted to working concentrations in the morning before beginning the protocol and were kept at 4°C throughout the day. Instead of tapping dry, wells were washed with 190 µL of wash buffer using a multichannel pipette ejecting waste into a bleach bath. Plates were read at 450 nm with 620 nm background subtraction on a Fluostar Optima platereader (BMG Labtech).

### Recombinant PrP

Recombinant PrP was expressed in *E. coli* and purified from inclusion bodies as described previously^47^ using a standard protocol^48^. Recombinant protein preparations were quantified by amino acid analysis (New England Peptide), purity assessed by Coomassie staining (Figure S3), and identity confirmed by LC/MS as described^47^ (Figure S3). All constructs were expressed in a pET-41a(+) vector. Human, mouse, Syrian hamster, and bank vole (M109) constructs were generous gifts from Byron Caughey, Andrew G. Hughson, and Lynne D. Raymond at Rocky Mountain Laboratories. Rat and cynomolgus constructs were produced by Genscript. Sequences (Table S3) were translated using ExPASy^49^ and aligned using Clustal Omega^50,51^ (Figure S3). Aliquots of 40 µL with 0.03% wt/vol CHAPS (Sigma C9426) were frozen at -80°C.

### Multiple reaction monitoring (MRM)

Multiple reaction monitoring was performed as described^12^. For rat brain analysis, one hemisphere of cortex and subcortex (without cerebellum or brainstem) were homogenized at 20% wt/vol in cold 0.2% wt/vol CHAPS, 1X PBS, and 1 tablet protease inhibitor (Roche cOmplete, Sigma 4693159001) per 10 mL, then diluted to 0.5% wt/vol homogenates in artificial CSF with final concentration of 0.03% wt/vol CHAPS. Rat brain and CSF were processed and analyzed in singlicate, with single residue ^15^N/^13^C-labeled heavy peptides as the reference standard and light:heavy (L/H) peak area ratio to estimate the concentration of PrP in each sample.

All human CSF samples were analyzed in duplicate with fully ^15^N-labeled HuPrP23-231 as the reference standard and light:^15^N (L/^15^N) peak area ratio used to calculate PrP concentration. Test-retest analysis utilized CSF pairs taken 2-4 months apart from 5 individuals deliberately selected to include two individuals with, and three without, CSF processing anomalies, as this affects test-retest reliability for PrP^11^. After confirmation of test-retest reliability (mean test-retest CV = 4.5% to 15.7% for the four peptides with technical replicate CV < 15%; Table S4), we proceeded to analyze only CSF samples from first study visits, rather than all study visits, for the remaining N=42 study participants. For N=29 replicates, the VVEQMCITQYER peptide was more abundant in met-ox than reduced form; for these replicates, the L/^15^N ratio was calculated using the met-ox form of both light and labeled protein. Any sample-peptide combination with technical replicate CV > 15% was excluded from downstream analysis.

### Tissue processing

Brains for analysis were homogenized at 10% wt/vol (e.g. 100 mg tissue + 1 mL buffer) except where otherwise indicated, in cold 0.2% wt/vol CHAPS, 1X PBS, and 1 tablet protease inhibitor (Roche cOmplete, Sigma 4693159001) per 10 mL, using 3x 40 second pulses on a Bertin MiniLys homogenizer in 7 mL tubes pre-loaded with zirconium oxide beads (Precellys KT039611307.7). Human CSF was collected as described^10,11^, rat CSF collection is detailed below; 0.03% wt/vol CHAPS (final concentration) was added to all CSF samples at the earliest possible moment after collection.

### Patient samples

CSF from asymptomatic *PRNP* mutation carriers and controls was collected through the Massachusetts General Hospital prion disease biomarker study and include samples previously described^11^. Participants were recruited through PrionRegistry.org, Rally (Mass General Brigham), Prion Alliance, CJD Foundation. Participants analyzed here had no mutation (N=21), E200K (N=12), D178N (N=6), P102L (N=4), or other *PRNP* mutation (N=4) and had each made 1-5 study visits (mean: 2.3) spanning a time period of up to ∼3.5 years. 0.03% CHAPS (final concentration) was added immediately after CSF sample collection.

Postmortem human brain samples were obtained from the Massachusetts Alzheimer’s Disease Research Center (MADRC). Samples were from *N*=5 control individuals without dementia, ages 50s-90s, postmortem interval 8-86 hours, *N*=4 male and *N*=1 female.

### Animals

All mice were C57BL/6. PrP ZH3 knockout mice^52^ on a C57BL/6J background were crossed to C57BL/6N animals. RML prions were intracerebrally inoculated at age 6-10 weeks as described^6^. Intracerebroventricular (ICV) ASO injections in mice were as described^6^ and were performed at age 7-10 weeks, except in prion-inoculated animals, where injections were at 60 days post-inoculation. Mice for brain regional studies were 12-14 weeks old. For mouse brains, whole hemisphere analyses included cerebella but excluded brainstem and olfactory bulbs. Ipsilateral (right) hemispheres were used for RNA analysis and contralateral (left) for protein analysis.

Rats were Sprague-Dawley males (age study; age 3-11 months) and females (pharmacodynamic study; body weight ∼300 g at study start). Rat CSF collection was performed under terminal anesthesia as follows. Occipital and nuchal areas were trimmed of hair and wiped with 70% ethanol. The heads of the rats were immobilized in stereotaxic instrument (ASI SAS-4100) while being maintained on 3% isoflurane and warmed on a heating pad (Physitemp HP-1M). The nose was rotated down 45° and held in this position with the nose bar of the stereotax. A 90° hemostatic forceps (Roboz RS-7291) was depressed against the skin to locate the space between the trapezii and the base of the skull, and a 27G butterfly needle (MedVet International 26709) was held in a custom stereotaxic needle holder and attached to a 1 mL syringe, then it was inserted through the nuchal skin by lowering the dorsal/ventral knob of the stereotactic instrument. The plunger of the syringe was withdrawn to create vacuum and then the needle lowered further, into the cisterna magna, until CSF began flowing into the butterfly tubing. When CSF flow ceased or blood was observed, the tubing was clamped with a hemostat and, if necessary, the tube was clipped at the meniscus of blood. The syringe was plunged to eject CSF into a low protein binding microcentrifuge tube (Eppendorf 022431081), and 3% wt/vol CHAPS stock solution was added at a 1:100 dilution to yield a final concentration of 0.03% CHAPS.

Cynomolgus macaque (N=3 male, N=3 female, age 2-4 years) tissue punches were obtained from tissue archived at -80°C from control animals treated with artificial CSF as part of previous ASO studies.

### Intracerebroventricular injections in rats

Rats were shaved and maintained at 3% isoflurane while being warmed with a heating pad (Physitemp HP-1M). They were placed in stereotaxic instrument (ASI Instruments, SAS-4100) with 27° atraumatic ear bars (ASI Instruments, EB-927), with the rat gas adapter riser set to -6 mm to set the lambda and bregma landmarks flat. The scalp was swabbed with betadine and ethanol and a 1.5 cm midline scalpel incision was made, centered between the nose and occipital ridge. Sterile cotton-tipped applicators were used to retract the subcutaneous and periosteal tissues. A sterile 1 mm x 33 mm drill bit (McMaster Carr, 5058N51) in a hanging-style handpiece (McMaster Carr, 4454A14) was positioned above bregma in a stereotactic handpiece holder (ASI Instruments, DH-1000) and then moved 1 mm caudal and 1.5 mm lateral. A bore hole was drilled at low speed and then a gastight 1710 small RN syringe (Hamilton 81030) was lowered through the skull hole, 3.7 mm from the surface of the brain into the lateral ventricle. 30 µL injection solution was ejected gradually over 10 seconds and the needle was retracted after 3 minutes. The incision was closed with 5-O monofilament suture (Ethilon 661G-RL) and rats recovered in their home cages.

### Statistics

All analyses utilized custom scripts in R 4.0.4. Statistical tests for each specific analysis are described throughout the text and figure legends. Tests are two-sided and P values are nominal except for GTEx analyses, which are Bonferroni-corrected for the number of tissues studied. All code and all raw data, except for potentially sensitive patient data from the clinical cohort, are available in a public git repository and can be used to reproduce the analyses herein: https://github.com/ericminikel/cns_prp_quant

### Study approval

Collection and analysis of human clinical samples were approved by the Partners Institutional Review Board (protocol #2017P000214). Animal studies were conducted under approved Institutional Animal Care and Use Committee protocols (Ionis Pharmaceuticals P-0273, Broad Institute 0162-05-17, and McLaughlin Research Institute 2020-DEC-75).

## Data Availability

Raw data and source code sufficient to reproduce the figures and analyses herein are available in the online git repository of this study with the exception of sensitive patient data, which are available upon reasonable request to the authors.

https://github.com/ericminikel/cns_prp_quant

## ACKNOWLEDGMENTS

This study was funded by Prion Alliance, CJD Foundation (the Michael H. Cole, Cheryl Molloy, José A. Piriz and Sonia E. Piriz, Jeffrey A. Smith, and Mercies in Disguise Memorial Grants), Ionis Pharmaceuticals (internal efforts and support to DEC), the Broad Institute (including direct philanthropic donations to Prions@Broad), the National Institutes of Health (R21 TR003040 to SEA), Ono Pharma Foundation, and an anonymous organization. We thank Brittany Ford and Adam Swayze for technical assistance.

## AUTHOR CONTRIBUTIONS

EVM and SMV conceived and designed the experiments. MM, HTZ, AGR, JEG, EK, JOM, PMD, TRC, CM, SWA, BAT, AM, MRM, and DEC performed the experiments. EVM, SMV, HTZ, JL, HBK, JM, DEC and SEA supervised the research. EVM performed statistical analyses and drafted the manuscript. All authors reviewed and approved the manuscript.

## SUPPLEMENTARY MATERIAL

### Development and evaluation of the cross-species PrP ELISA assay

Four commercially available antibodies with advertised species cross-reactivity were screened in all possible capture-detection configurations to identify suitable pairs for sandwich ELISA. This screen yielded four hits with promising signal-to-noise ratio (Figure S1A). All of these configurations proved dose-responsive and exhibited at least some cross-reactivity (Figure S1B-E). The EP1802Y capture and 8H4 detection configuration was selected as having the most similar dose-response curves for recombinant rat and human PrP (Figure S1E). An initial configuration of this assay was then validated for rat CSF (Appendix 3).

**Figure S1.**
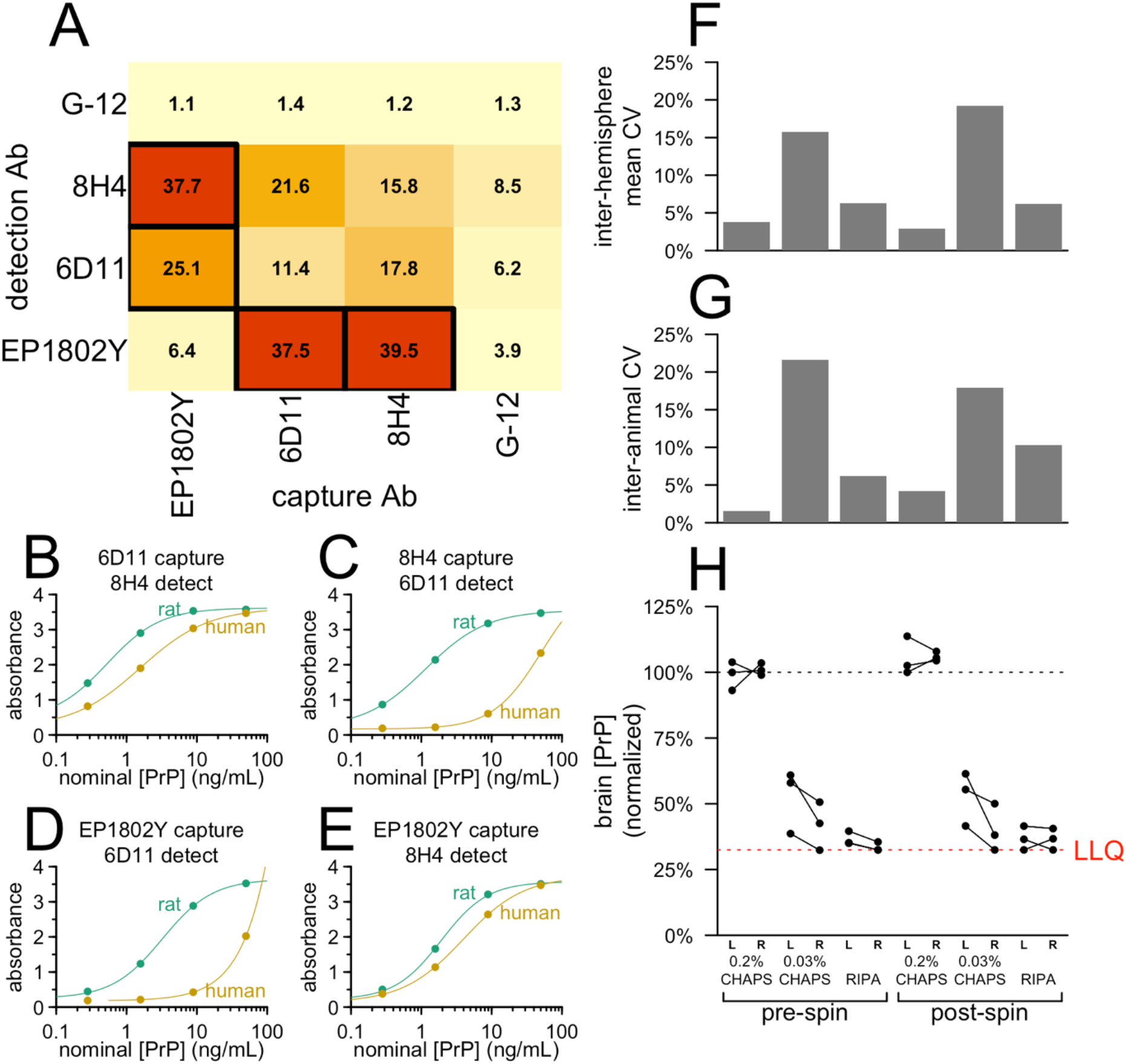
Development of the cross-species PrP ELISA. **A)** Signal-to-noise ratios (450 nm absorbance for 20 ng/mL vs. 0 ng/mL recombinant rat PrP) for screened antibody pairs. **B-E)** Dose-response curves for recombinant human and rat PrP for top four antibody pairs. **F)** Mean CVs comparing right vs. left brain hemispheres of the same animal, **G)** mean CVs between animals, and **H)** normalized response data for brains homogenized with the indicated detergents.

PrP in CSF exhibits enormous inter-individual variability if preanalytical variables are not properly controlled^10^, and we hypothesized the same might be true for PrP in brain tissue. We therefore sought to establish conditions for brain homogenization that would enable reliable PrP quantification. We hemisected frozen brains from wild-type mice, and for each animal, both right and left hemispheres were homogenized at 10% wt/vol in either 0.2% or 0.03% wt/vol CHAPS, or RIPA buffer (Pierce 89900, 25 mM Tris HCl pH 7.6, 150 mM NaCl, 1% NP-40, 1% sodium deoxycholate, 0.1% SDS). Homogenization in 0.2% CHAPS, just below the critical micelle concentration^53,54^, resulted in tight agreement of PrP concentration between hemispheres (mean CV = 3.8%, Figure S1F) and between animals (mean CV = 1.5%, Figure S1G), with >2x higher PrP recovery (Figure S1H) compared to 0.03% CHAPS or RIPA.

After establishing the final standard curve points and assay concentrations (see Methods and Appendices 1-2), we sought to characterize the assay’s performance and determine whether it is fit for purpose for measuring PrP in mouse brain tissue in preclinical drug discovery experiments. We prepared quality control (QC) samples using mouse brain homogenized at 10% wt/vol in 0.2% CHAPS (Table S1), intended to represent brains with 100%, ∼50%, 10%, and 0% wild-type levels of PrP (high, mid, low, and negative QCs respectively) and analyzed them at a final 1:200 dilution (1:20 dilution of 10% wt/vol homogenate). A non-GLP validation following FDA guidance^23^ determined a dynamic range of 0.05 to 5 ng/mL, with acceptable precision for both calibrators and QCs across this range, except for the low QC sample, which had a high inter-plate CV (32.7%; Table S1). We further conducted a stability assessment for common preanalytical perturbations (Table S2). In contrast with CSF^10^, brain homogenate did not disclose a decrease in PrP concentration upon transferring between plastic tubes (Table S2). Instead, the most important variable was time the brain homogenate spent at room temperature or 4°C, with apparent PrP concentration increasing by 29-56% after 4 hours at either temperature.

**Table S1.**
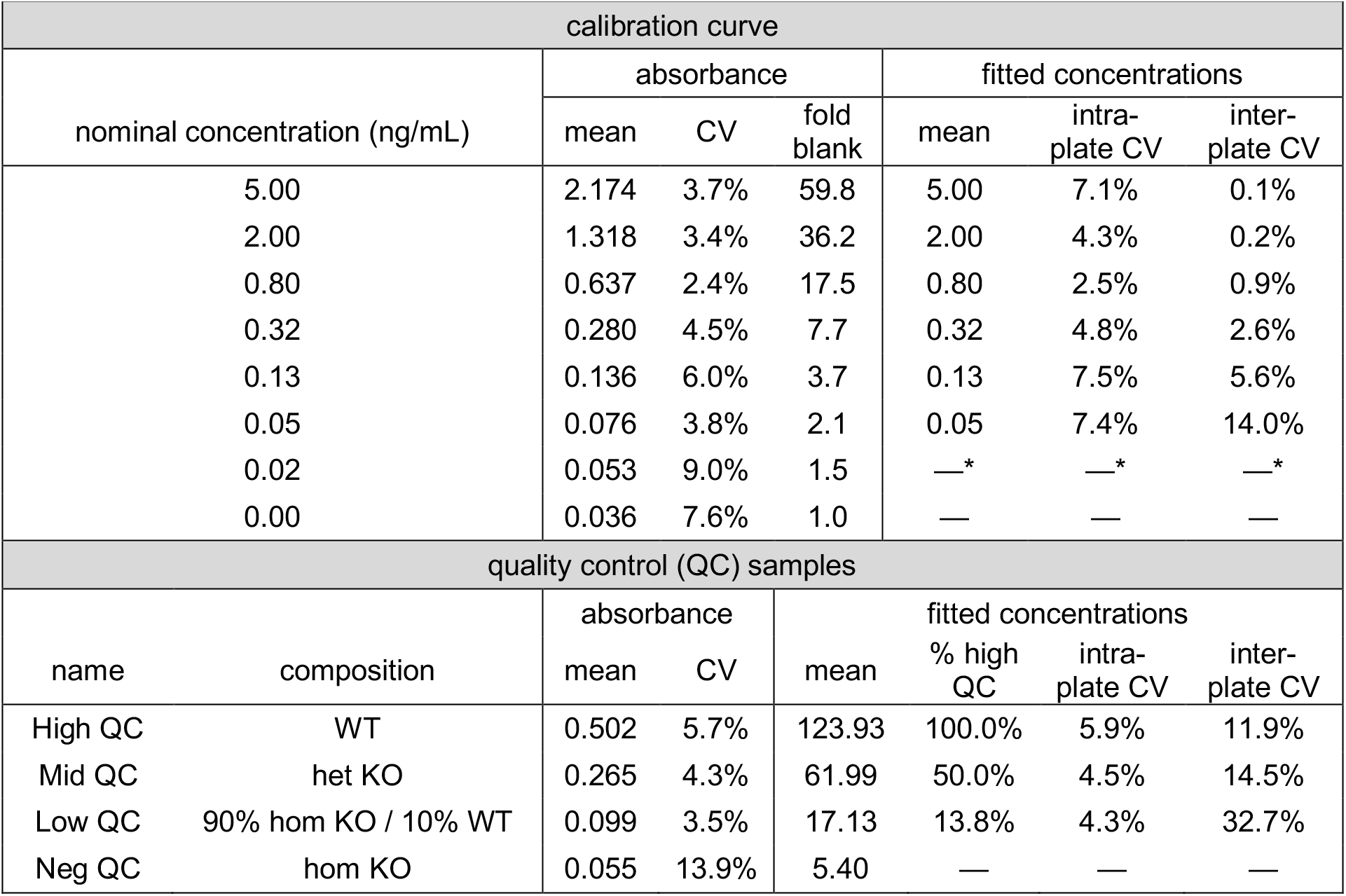
Performance of calibration curve and quality control samples in cross-species PrP ELISA. Inter-plate data are across seven validation plates; intra-plate data are from six replicates on one validation plate. For the analyses shown here, only standard curve points from 0.05 to 5.00 ng/mL were included in the four-point curve fit. *When the 0.02 ng/mL standard was included in the fit, its own mean backfit concentration was 0.01 ng/mL and its intra- and inter-plate CVs were 38.1% and 39.5% respectively.

**Table S2.** Stability assessment of mouse brain homogenate in cross-species ELISA. The indicated (n) number of aliquots of the same high and low PrP brain homogenate samples were subjected to a battery of conditions to determine mean apparent PrP concentration, coefficient of variation (CV) and absolute relative error (%RE).

| condition | high PrP (WT brain) |  |  |  | low PrP (90% KO / 10% WT) |  |  |  |
| --- | --- | --- | --- | --- | --- | --- | --- | --- |
|  | n | mean | CV | %RE | n | mean | CV | %RE |
| Freshly Thawed | 8 | 110.7 | 6% | — | 8 | 20.2 | 14% | — |
| Room Temp 4hrs | 4 | 161.3 | 2% | **46%** | 4 | 31.5 | 2% | **56%** |
| 4°C 4hrs | 4 | 142.3 | 1% | **29%** | 4 | 26.1 | 10% | **29%** |
| Freeze Thaw 1 cycle | 4 | 117.5 | 4% | 6% | 4 | 24.3 | 12% | 20% |
| Freeze Thaw 2 cycles | 4 | 129.0 | 4% | 17% | 4 | 29.3 | 9% | **45%** |
| Transfer Plastic 1 cycle | 4 | 111.0 | 5% | 0% | 4 | 24.6 | 7% | 22% |
| Transfer Plastic 2 cycles | 4 | 117.3 | 6% | 6% | 4 | 25.3 | 8% | 25% |
| Transfer Plastic 3 cycles | 4 | 127.4 | 4% | 15% | 4 | 22.6 | 17% | 12% |

We sought to determine across what dilutions the assay might exhibit the property of parallelism, meaning that a sample plated at different dilutions results in the same dilution-adjusted concentration. The adjusted concentrations for all QCs rose at progressively weaker dilutions, even up to the lower limit of quantification of the assay (Figure S2A). However, the relative concentration of PrP in mid and low QC samples compared to the high QC remained constant regardless of dilution (Figure S2B). This suggested that while progressive dilution of brain homogenate into assay buffer changes the apparent concentration of PrP in this assay, progressive dilution of endogenous PrP into brain homogenate does not. This was confirmed by preparing a 7-point dilution series of wild-type brain into PrP knockout mouse brain, which resulted in a linear response at a 1:200 final dilution (Figure S2C). Thus, this assay exhibits a linear response to PrP concentration in brain tissue, provided that brain samples to be compared are plated at the same dilution into assay buffer. For three control human CSF samples, however, parallelism was observed over dilutions from 1:5 to 1:80 (Figure S2D), in agreement with findings from a commercial PrP ELISA kit^10^. Standard curves of five species’ recombinant PrP reacted identically in our assay, while a sixth species, Syrian hamster, exhibited ∼3-fold lower, but still dose-responsive, reactivity (Figure S2E; see Figure S3 and Supplemental Discussion). For *N*=64 human CSF samples analyzed by both cross-species PrP ELISA and the commercially available BetaPrion ELISA kit, the rank order of concentrations was closely preserved (rho = 0.84, Spearman’s correlation), while the absolute PrP concentration read out in cross-species PrP ELISA was ∼6-fold lower (Figure 2F; see “Discussion of assay validation” status below).

**Figure S2.**
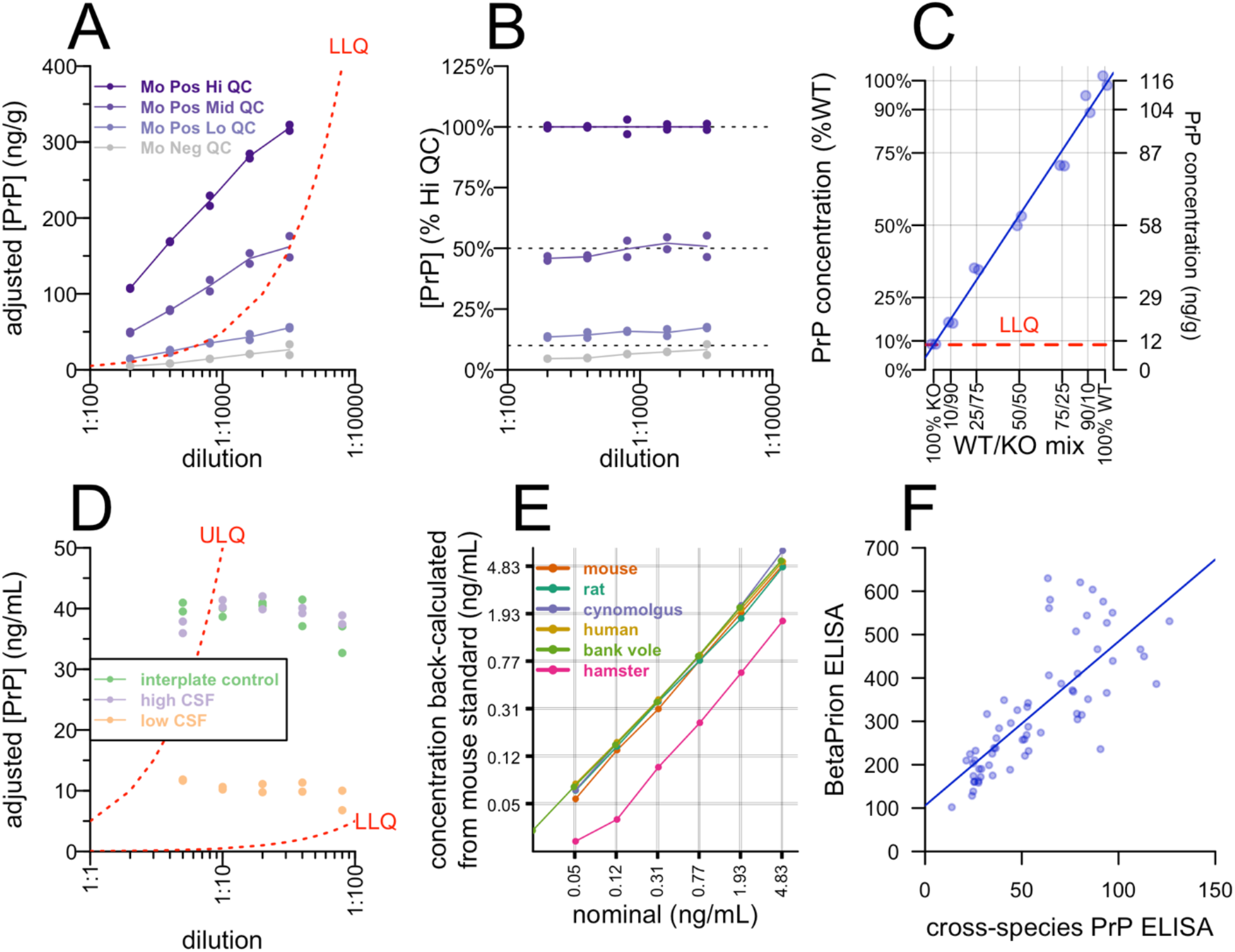
Parallelism, specificity, cross-reactivity, and comparison with BetaPrion ELISA. **A)** QC samples were plated at dilutions from 1:200 to 1:1,600, the y axis indicates the apparent concentration after adjusting for dilution. **B)** The data from A normalized to the adjusted concentration of the high QC. **C)** Specificity assessed by a dilution series of wild-type into knockout brain homogenate. The blue line is the best fit. **D)** Control human CSF samples were plated at dilutions from 1:5 to 1:80, y axis indicates dilution-adjusted concentration as in A. **E)** AAA-quantified recombinant PrP from six species was plated at nominal concentrations indicated by the x axis, the y axis shows the apparent concentrations back-fit to the mouse standard curve. **F)** Best fit between cross-species PrP ELISA and BetaPrion ELISA for N=64 human CSF samples from N=29 individuals analyzed by both methods.

**Table S3.**
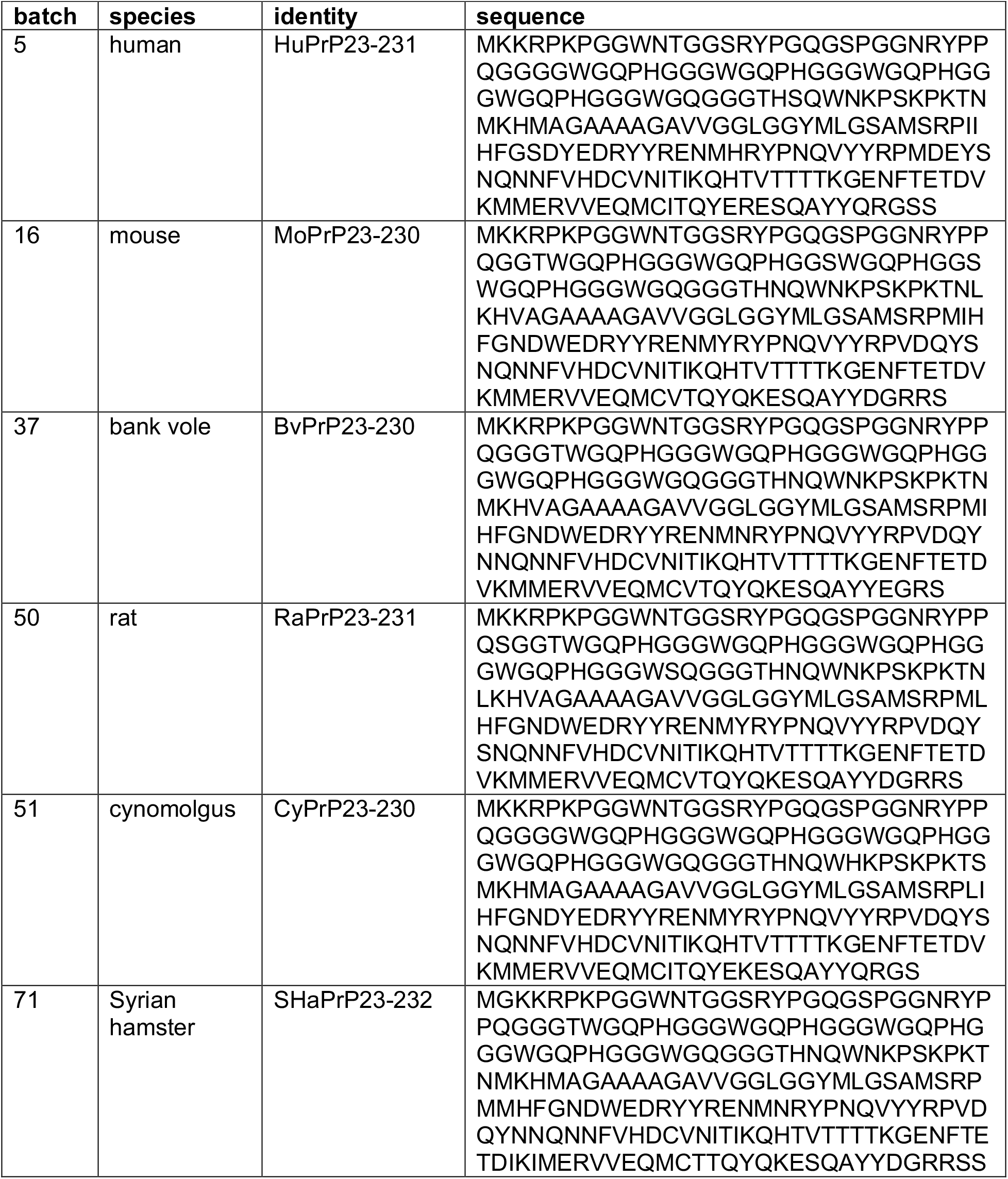
Recombinant PrP constructs. Note that N-terminal methionines in E. coli are expected to be cleaved when followed by G but not when followed by K^55^, see Figure S3. The first K in each sequence corresponds to residue K23 in humans or its ortholog in other animals, the first residue after PrP’s signal peptide.

**Figure S3.**
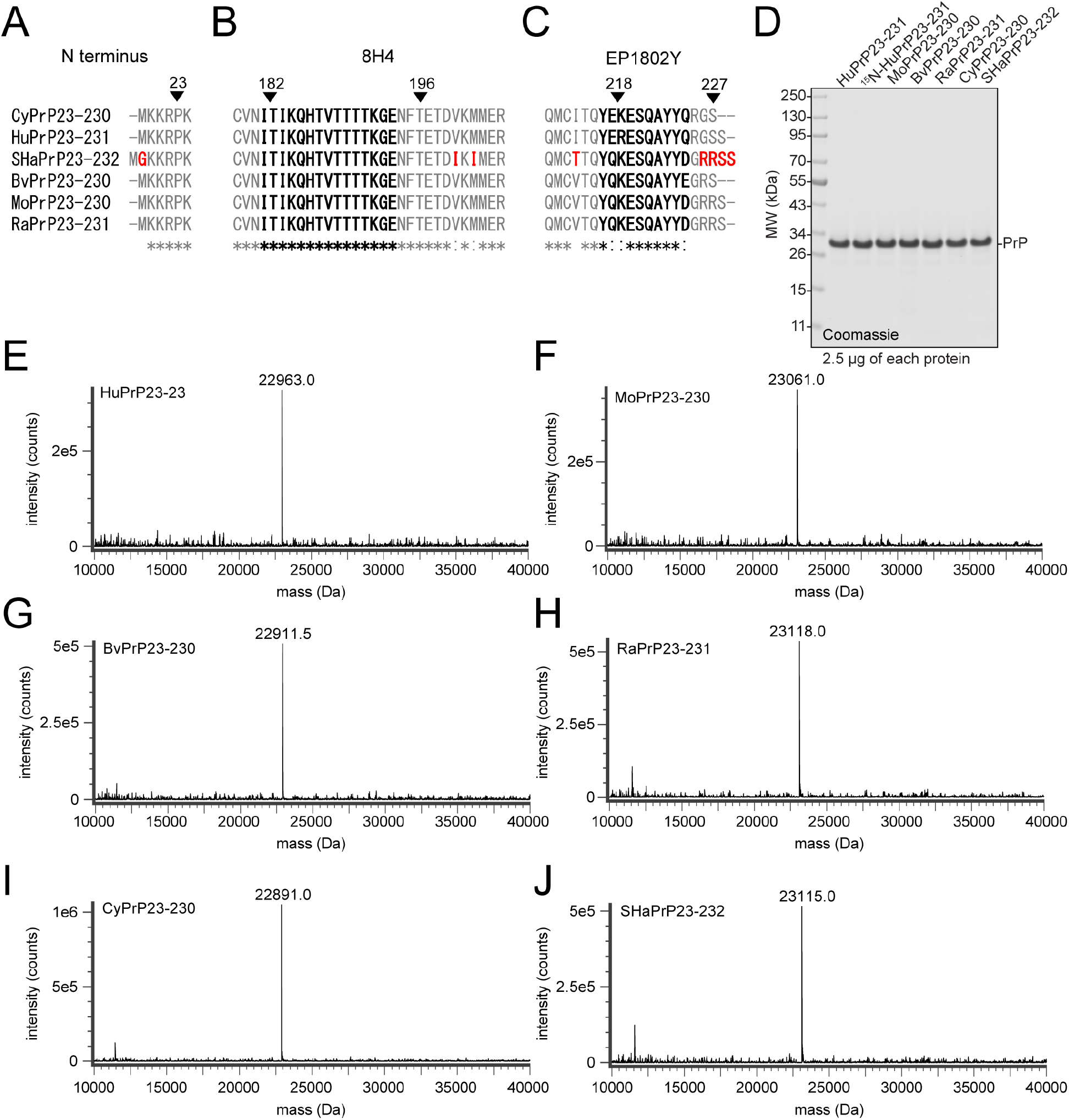
Epitope sequence, purity, and identity of recombinants. A-C) Multiple alignment of vector sequences at the N and C termini and reported antibody epitopes^20–22^, translated using ExPASy^49^ and aligned with Clustal Omega^50,51^. Residues reported to be part of the 8H4 and EP1802Y epitopes are in bold, and residues unique to the Syrian hamster construct are highlighted in red. D) Coomassie-stained SDS-PAGE of the six recombinant batches used as standards in the ELISA assay, plus the ^15^N-labeled HuPrP used as the standard in the MRM assay. E-J) Deconvoluted charge envelope of each recombinant standard run in intact protein LC-MS.

There are several possible explanations for the reduced reactivity observed for Syrian hamster PrP. The N terminus of our other five constructs contain a retained N-terminal methionine^12^, while the Syrian hamster construct contains a cleaved^55^ N-terminal methionine followed by a retained glycine (Figure S3A, red). The 8H4 antibody^20^ has been found nonreactive for squirrel monkey PrP, which contains an I182V substitution (human codon numbering; CNVN<u>V</u>TIKQ), as well as for the human mutations H187R and E196K^21^, suggesting its epitope spans from at least residue 182 to 196. These residues are invariant among the six species studied here (Figure S3B, bold). Syrian hamsters do harbor V203I and M205I substitutions (TETDIKIMERV) not found in any other species considered here (Figure S3B, red), though in order for these to affect 8H4 binding, the epitope would have to be discontinuous, as our MRM data indicate that our ELISA assay shows undiminished activity for PrP with the E200K mutation. Mutation scanning showed that the EP1802Y epitope was disrupted by mutations from residues 218 to 227 (human codon numbering)^22^. Syrian hamster PrP in this span is identical to both rat and mouse PrP (Figure S3C, bold), however it does harbor a nearby I215T substitution not seen in any other species here (Figure S3C, red). Finally, our Syrian hamster construct contains one additional residue of C-terminal sequence present in the other species’ genomes but not included in the recombinant constructs used here.

Although characteristics of this protein looked similar to the other batches employed here (Figure S3), we also considered technical explanations for the reduced reactivity of our Syrian hamster recombinant PrP. However, its elution curve was typical (Figure S4A), high purity by Coomassie (Figure S4B) was confirmed by size exclusion chromatography (Figure S4C), and identity was confirmed by LC/MS (Figure S4D). Despite all this, the lower reactivity compared to mouse PrP replicated identically across two plates (Figure S4E-F).

**Figure S4.**
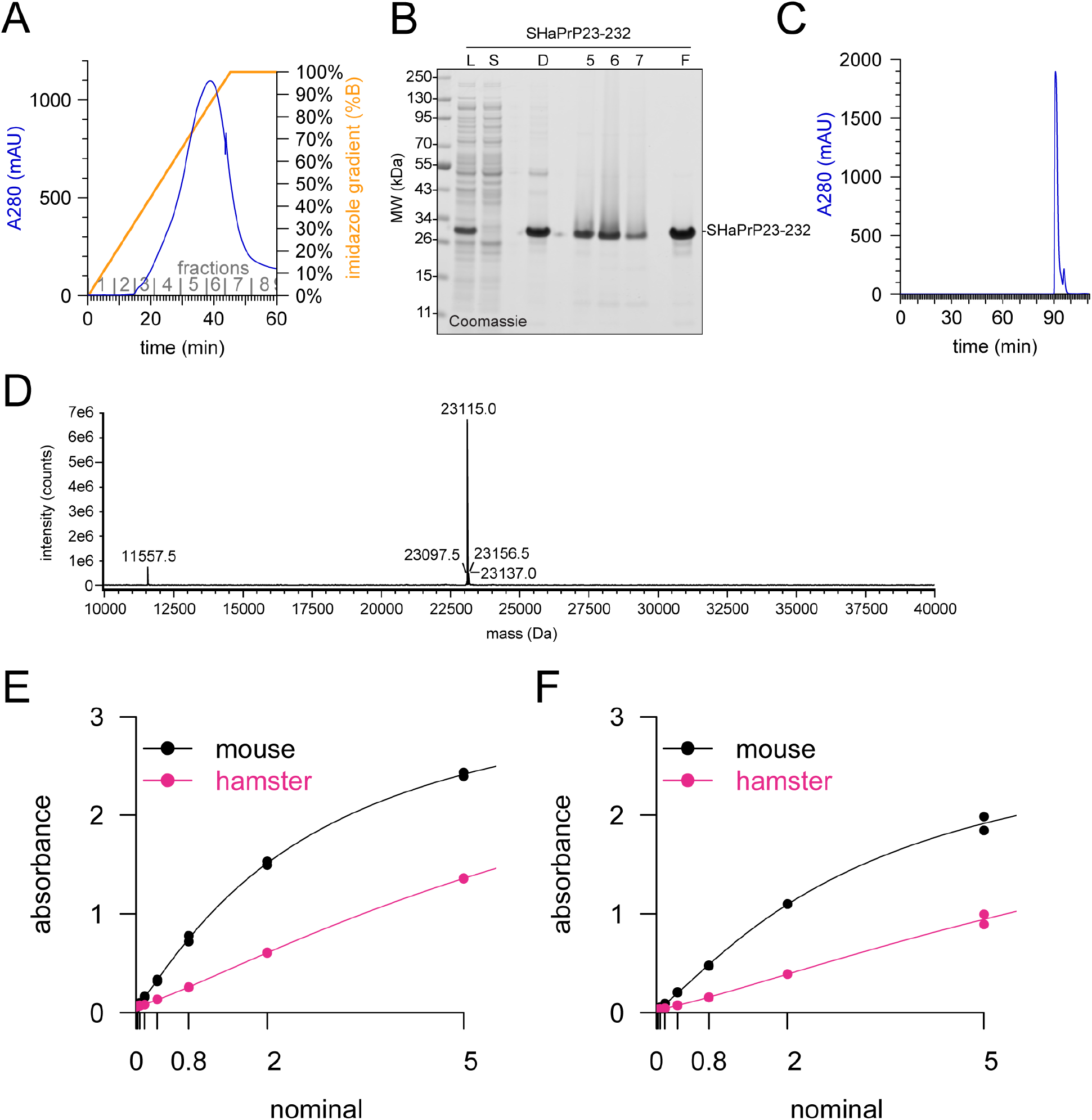
Hamster PrP purification and characterization. Figure S2. SHaPrP23-232 purification and characterization. A) AKTA UV chromatogram of IMAC elution. B) Coomassie-stained SDS-PAGE of fractions from the purification of SHaPrP23-232. L, whole-cell lysate (diluted 1:20); S, soluble fraction (diluted 1:20); D, guanidinium denatured protein (diluted 1:20); 5-7, AKTA IMAC elution fractions; F, final SHaPrP23-232 sample used as an ELISA standard. C) SEC UV absorbance chromatogram. D) Deconvoluted charge envelope of SEC purified SHaPrP23-232 from intact protein LC-MS. The mass of 23115.0 Da corresponds to SHaPrP23-232 without the N-terminal methionine, and with the intramolecular disulfide bond in the oxidized state. E-F) Raw calibration curves for mouse and hamster PrP run on two separate ELISA plates.

### Discussion of assay validation status

Bioanalytical methods used in drug development should be “fit for purpose,” with standards and expectations differing depending on the intended use case^23^. The data presented here indicate that our cross-species PrP ELISA is suitable for quantifying target engagement of PrP-lowering therapeutics in mouse brain tissue, with certain caveats. Preanalytical variables — particularly time spent above freezing — must be properly controlled, samples are best compared at the same dilution, and inter-plate variability at the lower end of the dynamic range may be higher than desired, leading to a need for within-plate comparisons or additional technical replicates. PrP in brain homogenate, unlike CSF, does not appear highly sensitive to plastic exposure, perhaps because the high protein, lipid, and detergent content mitigate sticking. Surprisingly, for reasons not yet understood, measurable PrP in brain homogenate does appear to rise with increased time spent above freezing. Based on recombinant PrP binding curves, the assay appears applicable across at least six species of interest for prion research, although we did not perform full validation for all of them. Our data also support analysis of CSF in this assay, though we did not perform full validation in the final assay configuration for this matrix. Importantly, our assay uses a frozen recombinant PrP calibrator curve quantified by amino acid analysis (AAA). The one commercially available PrP ELISA, BetaPrion, uses lyophilized calibrators which appear to have PrP concentrations substantially lower than advertised^10^, which limits that assay’s capacity for absolute quantification of PrP (Dr. Ashutosh Rao, FDA, Oct 31, 2019). Our assay may be suitable for quantification of PrP in human CSF in a clinical trial setting, but because we are not a GLP laboratory, we did not pursue a formal validation for this use case. One important limitation is that the manufacturer (Abcam) recommends short-term storage at +4°C for the EP1802Y antibody, whereas long-term banking of a single lot of antibody at -80°C would be desirable for long-term analysis of clinical trial samples. We did not assess stability of either of our antibodies at -80°C. Finally, while we demonstrated target engagement of ASOs in prion-infected animals, we have not investigated whether our assay exhibits equal reactivity to PrP^Sc^ as it does to PrP^C^. Some PrP antibodies, including 8H4, have been reported to exhibit diminished reactivity for PrP^Sc^ depending upon both the prion strain and the capture antibody employed^56^.

### Quality control of PrP MRM

Among the five short-term test-retest CSF pairs analyzed, two peptides had high CVs (>30%), but these were peptides that also had high technical replicate CVs (>15%) among these samples (Table S4), perhaps because overall recovery (both of light and ^15^N-labeled peptides) was relatively low. For the four peptides with low technical replicate CVs, test-retest CV was also low, supporting the analysis of just one CSF sample from each individual in Figure 3.

**Table S4.**
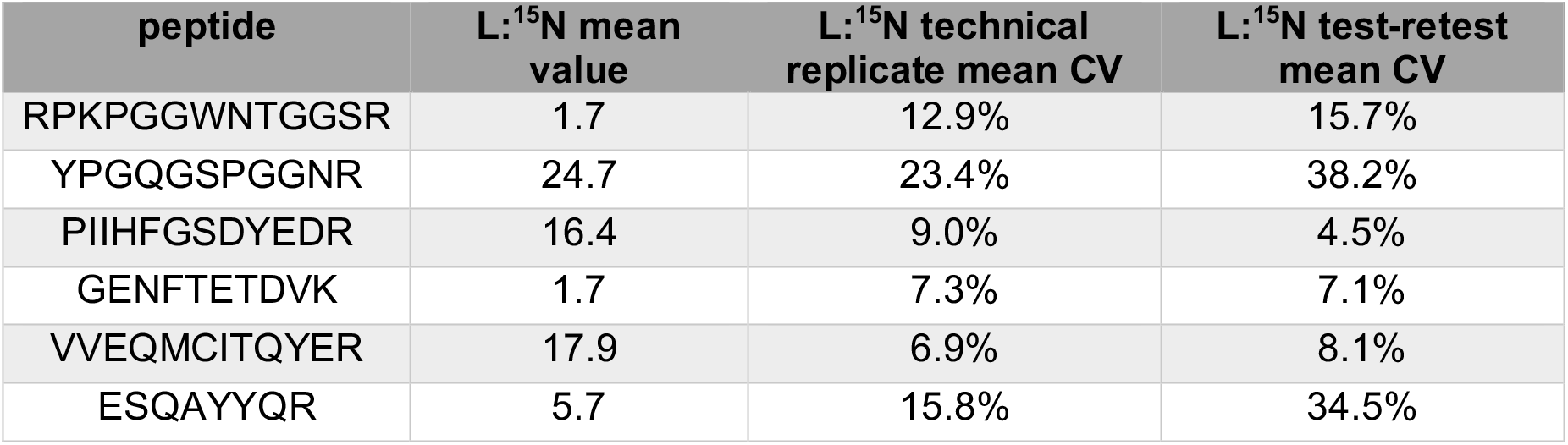
Performance of peptides in MRM on human CSF. For human sequence-matched peptides, we spiked fully ^15^N-labeled protein and used L:^15^N ratio as the assay readout. L:^15^N mean value and technical replicate mean CV are for all human CSF samples analyzed; test-retest mean CV is for the five test-retest pairs analyzed.

### Common variants in *PRNP*

We possessed only a small sample size of carefully handled CSF samples, and lacked genome-wide SNP data to control for population stratification. Nonetheless, in the interest of thoroughness, we chose to ask whether genotypes at two common *PRNP* variants with high prior probabilities for association with PrP expression showed any obvious correlation with CSF PrP concentration.

The coding variant rs1799990 (M129V) has dramatic effects on prion disease risk, duration, age of onset, clinical presentation, and histopathology across many subtypes of sporadic, acquired, and genetic prion disease^15^. For example, the heterozygous genotype is strongly protective against sporadic CJD in a genotypic model (OR = 0.39, *P* = 1e-135)^33^. It is the lead SNP for an eQTL for *PRNP* in several peripheral tissues but not in any brain region (Figure S5A). Our cohort contained only one VV individual, and there was no significant difference between CSF PrP in MM and MV individuals, whether all individuals or only mutation-negative controls were included (*P* = 0.06 or *P*=0.18, Kolmogorov-Smirnov test; Figure S5B).

Non-coding variant rs17327121, located 72 kb upstream of *PRNP*, is the lead SNP for an eQTL in cerebellum and cerebellar hemisphere, with no evidence of association with *PRNP* expression in any other brain region (Figure S5C). This SNP has not been reported to associate with prion disease risk, although neither it nor any SNP in tight linkage disequilibrium (r^2^ > 0.5 in CEU, computed using LDlink^57^) was genotyped or imputed in the largest sporadic CJD GWAS to date. None of the pairwise differences in CSF PrP between genotypes were significant (*P* > 0.2 for all pairs, Kolmogorov-Smirnov test; Figure S4D).

**Figure S5.**
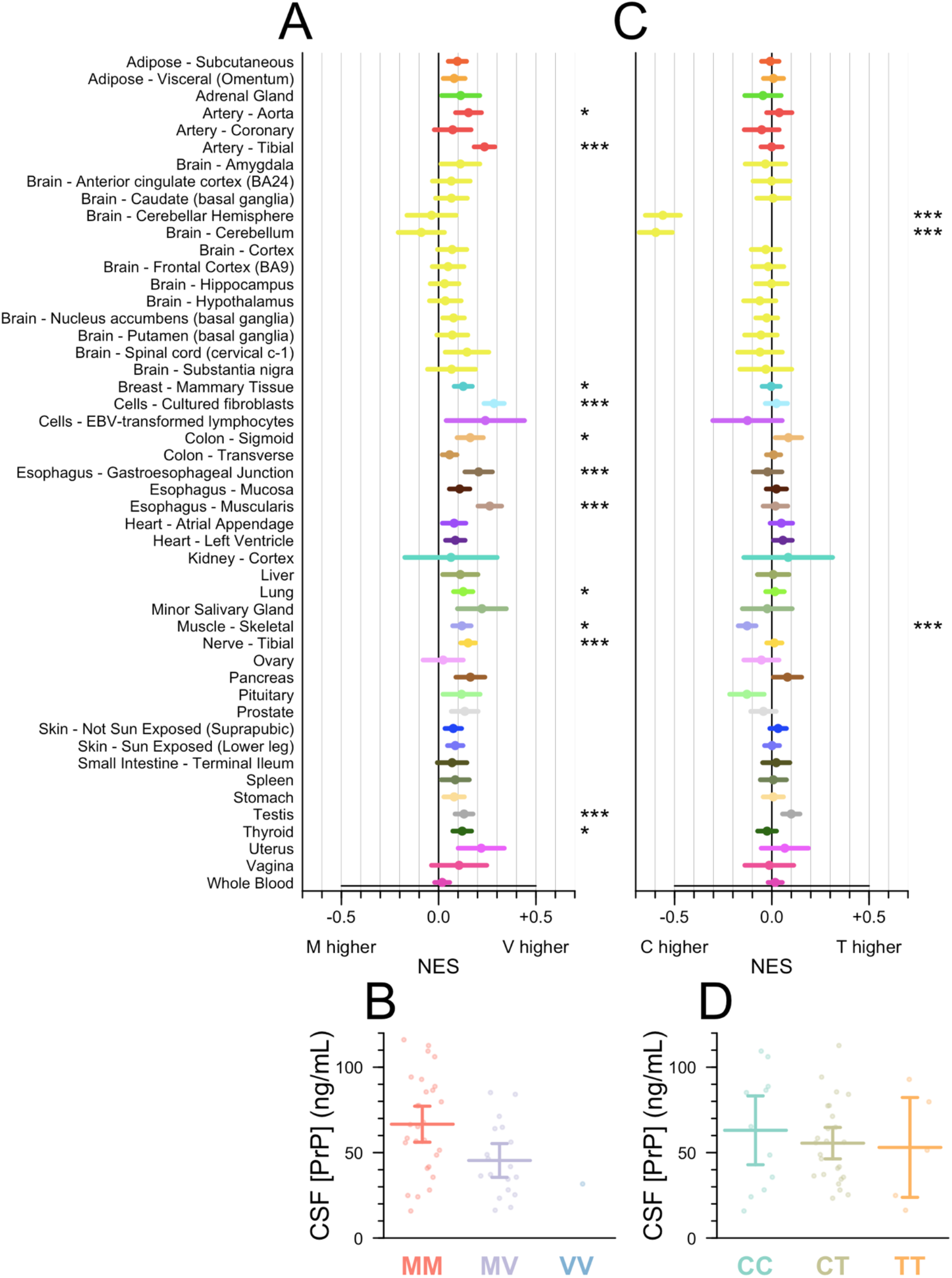
Common PRNP SNPs and CSF PrP. **A)** PRNP multi-tissue eQTL data for rs1799990 reproduced from the GTEx browser (gtexportal.org). Positions to the right of the zero indicate that the 129V haplotype is associated with higher PRNP RNA expression in certain tissues than the reference 129M haplotype. The x axis is normalized effect size (NES), which is performed on normalized expression values with no direct biological interpretation^26^. Empirical thresholds for significance^26^ in GTEx v8 vary by tissue down to 1e-5; symbols displayed here are as follows: * P < 1e-5, ** P < 1e-6, *** P < 1e-7. B) rs1799990 genotype and CSF PrP for all individuals in our MGH cohort. C) As panel A but for rs17327121. Positions to the left of the zero indicate that the reference allele, C, is associated with higher expression in cerebellum and cerebellar hemisphere than the alternate allele, T. D) rs17327121 genotype and CSF PrP in our MGH cohort.

### References

Citation numbers refer to references in the main text.

## Appendix 1. Full assay protocol

| Abbreviation | Name |
| --- | --- |
| Ab | Antibody |
| BSA | Bovine Serum Albumin |
| CHAPS | 3-[(3-Cholamidopropyl)dimethylammonio]-1-propanesulfonate hydrate |
| CSF | Cerebrospinal fluid |
| CV | Coefficient of variation |
| HRP | Horseradish Peroxidase |
| LLQ | Lower Limit of Quantification |
| OD | Optical Density |
| PBS | Phosphate Buffered Saline |
| PrP | Prion Protein |
| QC | Quality Control |
| RE | Relative Error |
| rPrP | Recombinant Prion Protein |
| RT | Ambient Room Temperature |
| SD | Standard Deviation |
| TMB | 3,3',5,5'-Tetramethylbenzidine |
| ULQ | Upper Limit of Quantification |

### Reagents

| Name | Manufacturer | Catalog # |
| --- | --- | --- |
| Zeba Spin Desalting Columns | Thermo Scientific | 89889 |
| EZ-Link Sulfo-NHS-SS-Biotin, No-Weigh format, 1 mg | Thermo Scientific | A39258 |
| 1X PBS | Broad Institute | N/A |
| Pierce BCA Protein Assay Kit | Thermo Scientific | 23225 |
| Anti PrP Ab 8H4 | Abcam | ab61409 |

### Equipment

| Name | Manufacturer | Model # |
| --- | --- | --- |
| 37°C Incubator | Any | Any |
| NanoDrop | Thermo Scientific | NanoDrop 8000 |
| SpectraMax M5 Plate Reader | Molecular Devices, Inc. |  |
| Standard Orbital Shaker | VWR | 1000 |

### Biotinylation of 8H4 Antibody

#### Solution Preparations

1. Dilute 90 µg of 8H4 Ab (e.g. 50 µL of 1.8 mg/mL) with 1X PBS to bring to a total 200 µL.

#### Material Buffer Exchange

2. Remove the bottom closure on the Zeba column and place into a clean 15 mL conical tube. Keep the column upright and cap loosened.
3. Centrifuge the column device at 1000xG for 2 mins. Flow-through is discarded and the device was placed back into the same falcon tube.
4. 1 mL 1X PBS was added directly on top of the resin. The device is centrifuged at 1000 RCF for 2 mins and the flow-through was discarded. This step is repeated two more times for a total of 3 washes.
5. After the last wash step, the column is removed from the conical tube. Keeping the column upright, the bottom of the column is blotted off with a Kimwipe and is transferred to a clean 15 mL falcon tube.
6. 200 µL of 8H4 Ab is applied directly on top of the resin. After 1 min, 40 µL of 1X PBS is applied as a stacker.
7. The device is centrifuged at 1000xG for 2 mins. The column is discarded and the flow-through is kept on ice. The volume collected from the device is measured using a pipette and recorded.

#### Biotinylation

8. 180 µL of cold Milli-Q water is added into a microtube of 1 mg of NHS-SS-Biotin to prepare a 10mMol Biotin stock solution. The contents are mixed with a pipette and then mini-centrifuged to bring the solution down.
9. **See note for calculations** 14.6 µL of 10mM Biotin stock solution is added into the 8H4 Ab solution and mixed with a pipette.
10. The biotinylated 8H4 Ab solution is covered in foil and placed on the plate shaker for 30 mins at the setting “4” (∼127 rpm).

#### Purification of Conjugated Protein

11. Remove the bottom closure on a new Zeba column and place into a clean 15 mL falcon tube. The column is kept upright and the cap loosened.
12. Following similar steps in the *Material Buffer Exchange* section, centrifuge the column device at 1000xG for 2 mins. The flow-through is discarded and the device was placed back into the same falcon tube.
13. 1 mL 1X PBS is added directly on top of the resin. The device is centrifuged at 1000xG for 2 mins and the flow-through was discarded. This step is repeated two more times for a total of 3 washes.
14. After the last wash step, the column is removed from the falcon tube. Keeping the column upright, the bottom of the column is blotted off with a Kimwipe and was transferred to a clean 15 mL falcon tube.
15. The biotinylated 8H4 Ab is applied directly on top of the resin. After 1 min, 40 µL of 1X PBS is applied as a stacker.
16. The device is centrifuged at 1000xG for 2 mins. The column is discarded and the flow-through is kept on ice.
17. The purified biotinylated 8H4 Ab solution is transferred into a clean 1.5 mL microtube, covered with foil and placed in the 4°C fridge. The final volume collected is measured using a pipette and recorded.
18. Use NanoDrop (Protein IgG concentration setting) to determine the concentration of the Ab. *Note: BCA can be used as an alternative to NanoDrop*.

### **Note

#### Calculations for Biotinylation

1. Calculate the concentration (mM) of the Sulfo-NHS-SS-Biotin to add to the reaction in order to obtain a specific molar excess. Typical challenge ratio is 20 Biotin: 1 molecule of protein for a 20 molar excess. The 8H4 Ab has a concentration of 1.8mg/mL in 50µL solution. Antibodies in general are ∼150 kDa or 150,000 mg/mmol. Equation used:

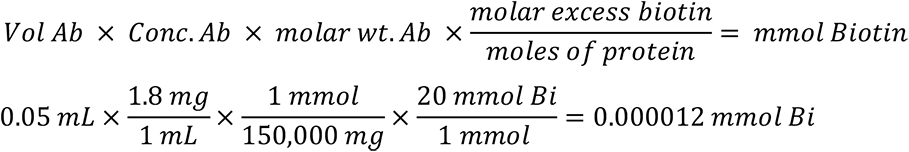
2. To calculate the volume (in µL) of 10 mM Sulfo-NHS-SS-Biotin to add to the labeling reaction, where MW Biotin = 906.7 mg/mmol:

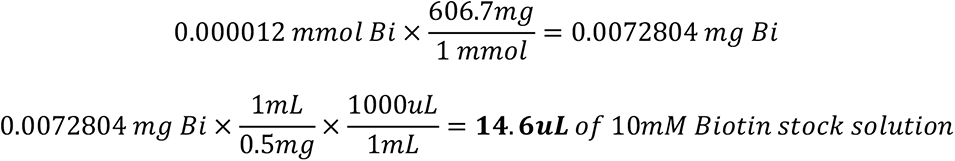

### Cross-Species PrP ELISA

#### Critical Equipment

| Description | Manufacturer | Model Number | Broad ID |
| --- | --- | --- | --- |
| SpectraMax M5 Plate Reader | Molecular Devices, Inc. |  | 101058 |

#### Critical materials, reagents, and supplies

| Name | Manufacturer | Model # |
| --- | --- | --- |
| Anti-PrP Ab EP1802Y | Abcam | ab52604 |
| Biotin-8H4 detection antibody | Broad Institute | N/A |
| Recombinant mouse prion protein | Broad Institute | Mo PrP16 |

#### General materials, reagents, and supplies

| Name | Manufacturer | Catalog # |
| --- | --- | --- |
| TMB substrate | Cell Signaling Technology | 7004P4 |
| Stop solution | Cell Signaling Technology | 7002L |
| CHAPS hydrate | Sigma | C9426 |
| Milli-Q water | Millipore | N/A |
| Pierce High Sensitivity Streptavidin-HRP | Thermo Scientific | 21130 |
| 96 Well Flat- Bottom Immuno Plate, MaxiSorp | Thermo Fisher Scientific | 439454 |
| 0.22µm vacuum filter system | Corning | CLS431098 |
| Bovine Serum Albumin | SeraCare Life Sciences | 19K15A0018 |
| 1X PBS CSHL, pH 7.4 | Broad Institute SQM | N/A |
| 10% Tween-20 solution | Teknova | T0710 |
| Seal, Clear Adhesive MicroAmp Film | Life Technologies | 4306311 |

#### Reagent Preparation

- <u>Wash buffer: 1X PBS with 0.1% Tween-20</u> Dilute 10% Tween-20 to 0.1% in 1X PBS. Example: 990mL 1X PBS + 10mL 10% Tween-20. Store at RT for up to 2 months
- <u>Assay buffer: 1X PBS with 5% BSA and 0.05% Tween-20</u> Dilute the required amount of BSA and 10% Tween-20 in 1X PBS. Mix thoroughly. Example: 25 g BSA + ∼400mL 1X PBS + 2.5 mL 10% Tween-20. Add 1X PBS to a final volume of 500mL. Filter through a 0.22 µm vacuum filter. Store at 4°C for up to 1 month.
- <u>Standards</u> Prepare high standard (Std01) by diluting stock MoPrP16 to 5ng/mL is assay buffer. Make 6 serial dilutions to produce the concentrations 2, 0.8, 0.32, 0.128, 0.0512, and 0.02048 ng/mL (Std02-07). The low standard (Std08) is neat assay buffer. Make a standard curve fresh from frozen, undiluted rPrP stock every time.
- <u>QC Samples</u> The QC samples used are: Mo Pos Hi QC, Mo Pos Mid QC, Mo Pos Lo QC, and Mo Neg QC. The QCs are stored at -80°C and are in 40 µL aliquots.

#### Procedure

1. Prepare capture Ab solution by diluting capture antibody EP1802Y to 2.0 µg/mL in PBS. Vortex briefly to mix. Prepare enough Capture Ab solution to add 100 µL to each plate well plus a 10% excess. Seal the plate and store <u>overnight</u> at 4°C.
2. Wash plate 3x with 300 µL Wash buffer per well. Tap dry.
3. Block plate by adding 250 µL Assay buffer per well. Seal and incubate at RT for 1-3 hours.
4. Wash plate 3x with 300 µL Wash buffer per well. Tap dry.
5. While the plate is blocking, dilute standards, QCs, and samples in assay buffer and add 100 µL of each to the plate per plate map. Pipette up and down to mix. Seal and incubate at RT for 60-75 minutes.
6. Wash plate 3x with 300 µL Wash buffer per well. Tap dry.
7. Prepare detection Ab solution by diluting biotin-labeled 8H4 detection antibody to 0.25 µg/mL in assay buffer. Vortex briefly to mix. Prepare enough detection Ab solution to add 100 µL to each plate well plus a 10% excess. Seal the plate and incubate at RT for 60-75 minutes.
8. Wash plate 3x with 300 µL Wash buffer per well.
9. Prepare streptavidin-HRP solution by diluting streptavidin-HRP to 24.69 ng/mL in assay buffer. Vortex briefly to mix. Prepare enough Streptavidin-HRP solution to add 100 µL to each plate well plus a 10% excess. Seal and incubate at RT for 20-30 minutes. *(**Note: full 30 minutes recommended, otherwise the plate may not reach ∼0.8 OD in the 30-minute time from during the TMB incubation step.)*
10. Wash plate 3x with 300 µL Wash buffer per well
11. Add 100 µL per well of TMB to plate. TMB solution should come to RT before using. Cover and incubate at RT until Std01 (5ng/mL) reaches ∼0.8 OD. Pre-read plate at 605nm. If Std01 does not reach this OD within 30 minutes stop plate and read.
12. Add 100 µL per well of Stop solution to plate. Stop solution should come to RT before using. Mix well on plate reader briefly and read at 450nm and 630nm.

## Appendix 2. ELISA working checklist

<u>Day 1</u>

1. Incubate the plate with 100 µL/well of **2 µg/mL EP1802Y Ab**. Seal and store at 4°C overnight.

Day 2

1. Wash plate 3X with 300 µL/well of **wash buffer** and tap dry
2. Block by adding 250 µL/well of **assay buffer** to plate. Seal and incubate at RT for 1-3 hr on benchtop Start time: _______ Sealed: _______ → Stop time: _______
3. Prepare fresh standards from an aliquot of stock rPrP
4. Wash plate 3X with 300 µL/well of **wash buffer** and tap dry
5. Add 100 µL/well of **rPrP standards, mouse QCs, and samples** in duplicate. Seal and incubate at RT for 60-75 min. Start time: _______ Sealed: _______→ Stop time: _______
6. Wash plate 3X with 300 µL/well of **wash buffer** and tap dry
7. Add 100 µL/well of **0.25 µg/mL biotin-8H4 Ab solution**. Seal and incubate at RT for 60-75 mins. Start time: _______ Sealed: _______→ Stop time: _______
8. Wash plate 3X with 300 µL/well of **wash buffer** and tap dry
9. Add 100 µL/well of **24.69ng/mL streptavidin-HRP solution**. Seal and incubate at RT for 30 mins. Start time: _______ Sealed: _______→ Stop time: _______
10. Wash plate 3X with 300 µL/well of **wash buffer** and tap dry.
11. Add 100 µL/well of RT **TMB**. Cover and incubate at RT on benchtop until Std. 1 (5ng/mL) reaches ∼0.8 OD (pre-read at 605 nm) or 30 minutes max. Start time: _______ Covered: _______→ Stop time: _______
12. Add 100 µL/well of RT **Stop Solution**. Mix well on plate reader briefly and read at 450 nm and 630 nm.

## Appendix 3. GCLP validation results for rat CSF

*Note: This validation study was performed by Bioagilytix Boston prior to the assay being transferred to the Broad Institute. The streptavidin-HRP concentration and the recombinant PrP standard curve points differ from the final assay configuration used at the Broad Institute. The results summary is shown below; the SOP and full validation report are available in this study’s online GitHub repository*.

| Parameter | Expectation | Observed Performance |
| --- | --- | --- |
| Intra-Assay Precision | $CV \leq 20\%$ at VS-H, VS-M, VS-L | VS-H: 3.20%<br>VS-M: 5.12%<br>VS-L: 3.42% |
| Inter-Assay Precision | $CV \leq 30\%$ at VS-H, VS-M, VS-L | VS-H: 12.4%<br>VS-M: 16.6%<br>VS-L: 21.3% |
| Accuracy | $\pm 3$ SD from average back calculated result of VS-H, VS-M, VS-L | VS-H: 13.0 - 28.4 ng/mL<br>VS-M: 4.00 - 11.9 ng/mL<br>VS-L: 1.61 - 7.28 ng/mL |
| Limits of Quantitation | Lowest and highest standards with $CV$ and $RE \leq 25\%$ | Std01: $CV = 0.27\%$, $RE = 0.181\%$<br>Std07: $CV = 9.62\%$, $RE = 3.35\%$ |
| Parallelism | $CV \leq 30\%$ for all concentrations within the LOQ | Run05 (64 to 128-fold): 26.2%<br>Run06 (8 to 64-fold): 16.9%<br>Run07 (8 to 16-fold): 27.3% |
| 4°C Stability | $RE \leq 20\%$ for at least 2/3 of aliquots per lot of CSF tested | 100% of aliquots pass, $CV$ of -5.9 to 6.7% |
| RT Stability | $RE \leq 20\%$ for at least 2/3 of aliquots per lot of CSF tested | 67% of aliquots pass, $CV$ of -6.7 to -5.9%. Failing aliquot $CV = -29.4\%$ |
| Freeze Thaw Stability | $RE \leq 20\%$ for at least 2/3 of aliquots per lot of CSF tested | 100% of aliquots pass, $CV$ of 0 to 6.7% |

